# Comparing physical activity policy evaluations across 27 EU member states: Diverse indicators, inconsistent findings

**DOI:** 10.64898/2026.09.04.26361720

**Authors:** Sven Messing, Antonina Tcymbal, Leonie Birkholz, Adrian Bauman, Stephen Whiting, Colin O’Hehir, Catherine Woods

**Author notes:** <u>Corresponding author:</u> Dr. Sven Messing, University of Limerick, Physical Activity for Health Research Centre, Health Research Institute, Department of Physical Education and Sport Sciences, Castletroy, Limerick, V94 T9PX, Ireland,.

## Abstract

**Background:** Physical inactivity remains a major public health challenge, and policy action across sectors is needed to create environments that enable active lifestyles. Several tools have been developed to evaluate national physical activity policies, but it remains unclear whether they assess similar policy aspects or produce comparable results. This study compared evaluation results across the 27 EU member states and examined the indicators used by these tools.

**Methods:** Eight physical activity policy evaluation tools were included that (a) assess national physical activity policies and (b) had been applied in multiple countries. For five tools covering at least half of the EU member states, aggregate scores were extracted or calculated and converted into country rankings. A total of 202 indicators were coded and mapped by category (policy content, process, structure), and data type (audit, content, implementation, impact assessment).

**Results:** Country rankings varied considerably, with a mean difference of 13.0 positions between highest and lowest ranks. Some countries displayed consistent results (e.g., Austria, France, Germany), while others showed large discrepancies (e.g., Hungary, Spain). Indicator mapping revealed a focus on policy content (47.6%), followed by structure (21.6%) and process (13.7%) (6.2% other). Across tools, 20.3% of indicators audited policies, 26.0% assessed policy content, and 26.4% assessed policy implementation (16.3% other).

**Conclusion:** The systematic comparison of indicators across physical activity policy evaluation tools provided insights into differences in country rankings. Future evaluations should clearly communicate their purpose, identify synergies between tools, address gaps in policy evaluation, and strengthen the reproducibility of evaluation processes.

## Introduction

Physical inactivity is a public health problem, with 31.3% of adults and 81.0% of adolescents worldwide failing to achieve recommended levels of physical activity (Guthold et al., 2020; Strain et al., 2024). Even small increases in moderate-to-vigorous physical activity of just 5 minutes per day could reduce mortality and healthcare costs (Ding et al., 2016; Ekelund et al., 2026; WHO, 2022). However, progress is unlikely without “upstream” policy action, as programme-based “downstream” interventions alone cannot change the broader systems and environments that shape physical activity behaviours (Woods et al., 2022).

Consequently, the World Health Organisation (WHO) has set a target to reduce global physical inactivity by 15% by 2030, and emphasized the role of public policies in achieving this goal (WHO, 2018). This call to action is supported by evidence on the effectiveness of policy approaches in creating environments that support physical activity (Gelius et al., 2020). In the EU context, policy importance is also supported by EU Council Recommendations on promoting health-enhancing physical activity across sectors (Council of the European Union, 2013). To evaluate the implementation of these recommendations by EU member states, the European Commission developed a tool to enable regular data collection on 23 indicators. Similarly, WHO, non-governmental organisations and research groups have developed different tools to evaluate national physical activity policies (Vlad et al., 2023; WHO, 2022; Woods et al., 2022).

These tools are frequently used to assess and compare public policies for physical activity promotion at country level. For instance, the Global Observatory for Physical Activity (GoPA!) categorizes country’s policy landscape as “high”, “medium” or “low”, while the Active Healthy Kids Global Alliance (AHKGA) assigns grades for policies under its “government” indicator (Aubert et al., 2022; Ramirez Varela et al., 2025). Interestingly, the results of these policy evaluations seem to differ: For example, Croatia receives a “high” policy assessment in the GoPA! country card but a comparatively weak grade (D+) in the children-focused AHKGA report card (Pedisic et al., 2023; Ramirez Varela et al., 2025). Even though distinct purposes of the tools may explain these differences – with GoPA! focusing on adults and AHKGA on children and adolescents – these contrasting findings require further investigation. Previous studies have highlighted the issues of unclear relationships between these different tools and the use of different indicators (Tremblay et al., 2015). Understanding these differences is important because evaluation outputs shape how physical activity policy is understood by policymakers, practitioners and researchers. Previous reviews have shown that different tools have been developed for different purposes (Klepac Pogrmilovic, O’Sullivan, et al., 2019), and that government involvement varies considerably across tools (Messing et al., 2023). Data from some of these tools have been used to rank countries for comparison and advocacy purposes (Active Healthy Kids Global Alliance, 2022; AOK & dkfz, 2025). However, no previous study has systematically compared country rankings across physical activity policy evaluation tools, nor examined the indicators on which these tools are based.

To address this gap, our study analyses the results of physical activity policy evaluations across all 27 EU member states and maps the indicators used in international tools. We define physical activity policy as decisions, plans and actions that are enforced by governments which may directly or indirectly influence physical activity at population level (Lakerveld et al., 2020). As policy is a multidimensional concept (Dye, 1978), individual tools may operationalise it differently. We also adopt a broad view of evaluation that includes auditing, benchmarking and monitoring, in line with scholars from public policy research (Dahler-Larsen, 2012). In short, this study aims to answer two research questions: (1) To what extent are physical activity policy evaluation findings consistent across EU member states? (2) How can differences between tools and their findings be explained?

## Methods

### Selection of tools

Physical activity policy evaluation tools were identified through reference checks of existing systematic literature reviews (Klepac Pogrmilovic, O’Sullivan, et al., 2019; Messing et al., 2023) and an additional search. Two members of the research teams screened 28 potentially relevant tools, and selected those that fulfilled the following inclusion criteria: (a) the tool contains indicators or questions on public policies for physical activity promotion across relevant sectors, and (b) the tool has been used for data collection at the national level in more than one country. A flowchart of the selection process and a list of excluded tools are presented in appendix 1.

Eight tools were included (table 1).

**Table 1:** Included physical activity policy evaluation tools.

| **Abbreviation** | **Name of the tool** | **Purpose of the tool** | **Responsible organisation** |
| --- | --- | --- | --- |
| AHKGA Report Cards | Active Healthy Kids Global Alliance Report Card | Assess child and adolescent physical activity, related indicators, and key sources of influence (Aubert et al., 2022) | Active Healthy Kids Global Alliance |
| EU Monitoring Framework | EU Health-Enhancing Physical Activity (HEPA) Monitoring Framework | Monitor implementation of Council Recommendations in EU member states (Council of the European Union, 2013) | European Union |
| GoPA! | Global Observatory for Physical Activity Country Cards | Monitor progress in surveillance, research, and policy (Ramirez Varela et al., 2025) | Global Observatory for Physical Activity (GoPA!) |
| HEPA-PAT | Health-Enhancing Physical Activity Policy Audit Tool | Provide a comprehensive overview of the breadth of current physical activity policies (Bull et al., 2015) | Project-based |
| INTEGRATE-PA-Pol | Interaction between National and Local Government Levels | Assess collaboration between policy levels | Global Observatory for Physical Activity |
|  | in Development and Implementation of Physical Activity Policies Tool | (Resendiz et al., 2024) | (GoPA!) |
| MOVING | MOVING Physical Activity Policy Benchmarking Tool | Assess the strength of policy design, focus on adolescents (WCRF & NIPH, 2024) | World Cancer Research Fund (WCRF) |
| PA-EPI | Physical Activity Environment Policy Index | Assess the extent of policy implementation (Woods et al., 2022) | PA-EPI Network |
| WHO Monitoring Framework | WHO Global Action Plan on Physical Activity Monitoring Framework | Monitor implementation of Global Action Plan in WHO member states (WHO, 2022) | World Health Organisation (WHO) |

Data availability was checked for all 27 EU member states. For four tools, data were available for at least 25 EU member states (EU Monitoring Framework, GoPA!, MOVING, WHO Monitoring Framework). The AHKGA Report Cards had available data for 16 EU member states (Aubert et al., 2022), the HEPA-PAT for 11 EU member states (Kahlmeier, 2024), the PA-EPI for two EU member states (Heuvelman et al., 2025; Volf et al., 2023), and INTEGRATE-PA-Pol – which was developed in Latin America – for one EU member state (Pratt et al., 2025).

### Country ranking comparison

For the five tools with data available for at least half of the 27 EU member states, aggregate scores were extracted from scientific publications (Aubert et al., 2022; Whiting et al., 2021) or calculated using a predefined scoring procedure specific to each tool (appendix 2), such as summing indicators scored with ‘yes’ or converting categorical ratings to numerical values (Ramirez Varela et al., 2025; WCRF & NIPH, 2024; WHO, 2022). A second researcher independently calculated aggregate scores for a random sample of six included countries (>20%), and discrepancies were resolved through review of the original sources. Aggregate scores were converted into country rankings to facilitate comparison across tools. Rankings were considered more appropriate than raw scores because the tools use different scoring scales and reporting formats (e.g., grades, categories, or number of indicators). For each tool, rank 1 represents the country with the highest score; countries with the same score were assigned the same rank, and subsequent ranks were skipped.

To assess alignment across tools, the differences between highest and lowest rank were calculated for each country. In addition, the mean difference of country rankings was calculated first across all five tools, and then across the three tools with non-categorical data. The calculation of aggregate scores is described in detail in appendix 2.

### Mapping of indicators

For all eight tools, questionnaire items, benchmarks, good practice statements or indicators that guided data collection were identified; all referred to henceforth as ‘indicators’ to allow for comparison.

These indicators were entered into a spreadsheet, and compared based on two complementary classification systems (categories, types).

The development of categories was informed by the political science distinction between policy content (*policy*), political process (*politics*), and political structure (*polity*) (Rohe, 1994). Categories were refined and operationalised through an iterative process combining deductive application of existing concepts and inductive identification of additional aspects emerging from the data:

- *Policy content (specific):* Sector- or setting-specific indicators related to policy solutions for physical activity promotion. Codes were primarily based on ISPAH’s Eight Best Investments, including sport, health, and transport (Milton et al., 2021). Separate codes were assigned to school and childcare.
- *Policy content (general):* Content-related indicators not specific to a single sector or setting. Codes include key policy documents (across sectors), guidelines, policy comprehensiveness, and goals/actions/tasks.
- *Political process:* Indicators relating to agenda setting, policy formulation, decision-making / adoption, implementation, and evaluation (Howlett et al., 2009).
- *Political structure:* Indicators relating to the institutional context of policymaking. Codes include coordination mechanisms, funding, government structure, health in all policies, leadership, surveillance and monitoring systems, and workforce development.
- *Other:* Indicators not primarily focused on policy content, process or structure. Codes include behavioural and other indicators.

Types of data collected by tools were differentiated based on distinctions in the scientific literature and guidance from the Centers for Disease Control and Prevention (CDC, 2012; Klepac Pogrmilovic, O’Sullivan, et al., 2019):

- *Audit:* Indicators documenting the existence or characteristics of a policy without evaluating its quality, adequacy or effectiveness. Audits do not include rating, grading or judging a policy (Klepac Pogrmilovic, O’Sullivan, et al., 2019).
- *Content assessment:* Indicators evaluating the written content of policies, including core components, implementation requirements or the evidence base (CDC, 2012).
- *Implementation assessment:* Indicators evaluating policy implementation, including processes, capacities, facilitators and barriers (CDC, 2012).
- *Impact assessment:* Indicators evaluating policy outcomes and impacts, including short-, medium-, and long-term outcomes, effectiveness, costs or cost savings (CDC, 2012).
- *Other:* Indicators not primarily focused on the auditing or assessment types described above.

Based on these conceptual considerations, the first author developed a codebook. Two researchers independently coded all indicators (inter-rater agreement 75.5%). Conflicts were resolved in a consensus meeting. Further information can be found in appendix 3.

## Results

### Country ranking of EU member states

The country ranking is presented in table 2 and includes data from the EU Monitoring Framework, the WHO Monitoring Framework, GoPA!, MOVING, and the AHKGA Report Cards. The highest country ranks for each tool are shown in green, the medium ranks in yellow, and the lowest ranks in red.

**Table 2:** Country rankings for EU member states across five different tools.

| Country | AHKGA Report Cards (2022) | EU Monitoring Framework (2024) | GoPA! (2025) | MOVING (2023) | WHO Monitoring Framework (2022) |
| --- | --- | --- | --- | --- | --- |
| Austria | No data | 13 | 14 | 10 | 11 |
| Belgium | No data | 4 | 1 | 5 | 20 |
| Bulgaria | No data | 18 | 1 | 22 | 18 |
| Croatia | 13 | 8 | 1 | 20 | 15 |
| Cyprus | No data | 23 | 25 | No data | 27 |
| Czechia | 13 | 18 | 1 | 20 | 21 |
| Denmark | 2 | 8 | 14 | 2 | 1 |
| Estonia | 3 | 13 | 23 | 16 | 13 |
| Finland | 1 | 1 | 1 | 2 | 7 |
| France | 3 | 2 | 1 | 5 | 1 |
| Germany | INC | 2 | 1 | 2 | 5 |
| Greece | No data | 13 | 25 | 12 | 23 |
| Hungary | 3 | 4 | 1 | 15 | 26 |
| Ireland | 3 | 4 | 1 | 1 | 5 |
| Italy | No data | 20 | 23 | 18 | 14 |
| Latvia | No data | 23 | 25 | 22 | 15 |
| Lithuania | 10 | 13 | 14 | 5 | 1 |
| Luxembourg | No data | 20 | 14 | No data | 9 |
| Malta | No data | 23 | 14 | 24 | 24 |
| Netherlands | No data | 8 | 1 | 12 | 18 |
| Poland | 11 | 13 | 14 | 18 | 9 |
| Portugal | 3 | 8 | 1 | 5 | 7 |
| Romania | No data | 27 | 14 | 25 | 24 |
| Slovakia | 9 | 4 | 14 | 10 | 22 |
| Slovenia | 15 | 20 | 1 | 12 | 11 |
| **Spain** | 11 | 26 | 1 | 9 | 4 |
| **Sweden** | 3 | 8 | 14 | 16 | 17 |

*INC = Incomplete; Green = High country ranks; Yellow = Medium country ranks; Red = Low country ranks*

The mean difference between the highest and lowest country rank across all five tools is 13.0. The greatest variation in country ranking was observed for Hungary, Spain (both 25 ranks), Bulgaria (21 ranks), Czechia, and Estonia (both 20 ranks). The smallest differences were observed for Austria, Cyprus, France, Germany, and Ireland (all 4 ranks).

If tools with categorical data (AHKGA Report Cards, GoPA!) are excluded from this comparison due to their lower ability to differentiate between country ranks, the mean difference between the highest and lowest rank is 8.3. For the three remaining tools (EU Monitoring Framework, MOVING, WHO Monitoring Framework), country rankings differ most for Hungary, Spain (both 22 ranks), and Slovakia (18 ranks). The countries with the most consistent rankings across these three tools are Malta (1 rank) and Austria, Czechia, Estonia, Germany, Portugal and Romania (all 3 ranks). Country rankings are most consistent between the EU Monitoring Framework and MOVING (mean difference between highest and lowest rank = 4.6 ranks) and between the WHO Monitoring Framework and MOVING (4.8 ranks), in comparison to the differences between the EU and WHO Monitoring Frameworks (7.3 ranks).

### Mapping of indicators

To better understand why rankings differ, we mapped the 202 indicators that were identified across all eight tools; this is an average of 25.3 indicators per tool (range 4-45). More than half of these indicators collect data on the policy content (n=108; 53.5%): Seventy-seven of these indicators (33.9%) have a specific focus on the content of policies in relevant sectors or settings, e.g., sport, health, active transport, or schools, while 31 of these indicators (13.7%) have a more general focus, e.g. on the availability of key policy documents such as national action plans. Data on the political structure, e.g. mechanisms for intersectoral collaboration, funding, or monitoring and evaluation systems, are collected by 49 indicators (21.6%). The political process, e.g. agenda setting, policy formulation or decision-making, is the main focus of 31 indicators (13.7%). Finally, 14 indicators that do not fit into any of the above categories were identified as ‘other’ (6.2%).

Figure 1 illustrates the different focus of each tool on physical activity policy. Specific content-related data are most prevalent in MOVING (82.6% of this tool’s indicators), the WHO Monitoring Framework (75.9%), and the EU Monitoring Framework (60.9%), while GoPA! has a stronger focus on general content-related data (50.0%). Compared to the other tools, the indicators related to the political structure can more frequently be found in the AHKGA Report Cards (50.0%), GoPA (50.0%), the PA-EPI (42.2%) and the HEPA-PAT (37.8%). INTEGRATE-PA-Pol focuses primarily on the political process (60.0%).

**Figure 1:**
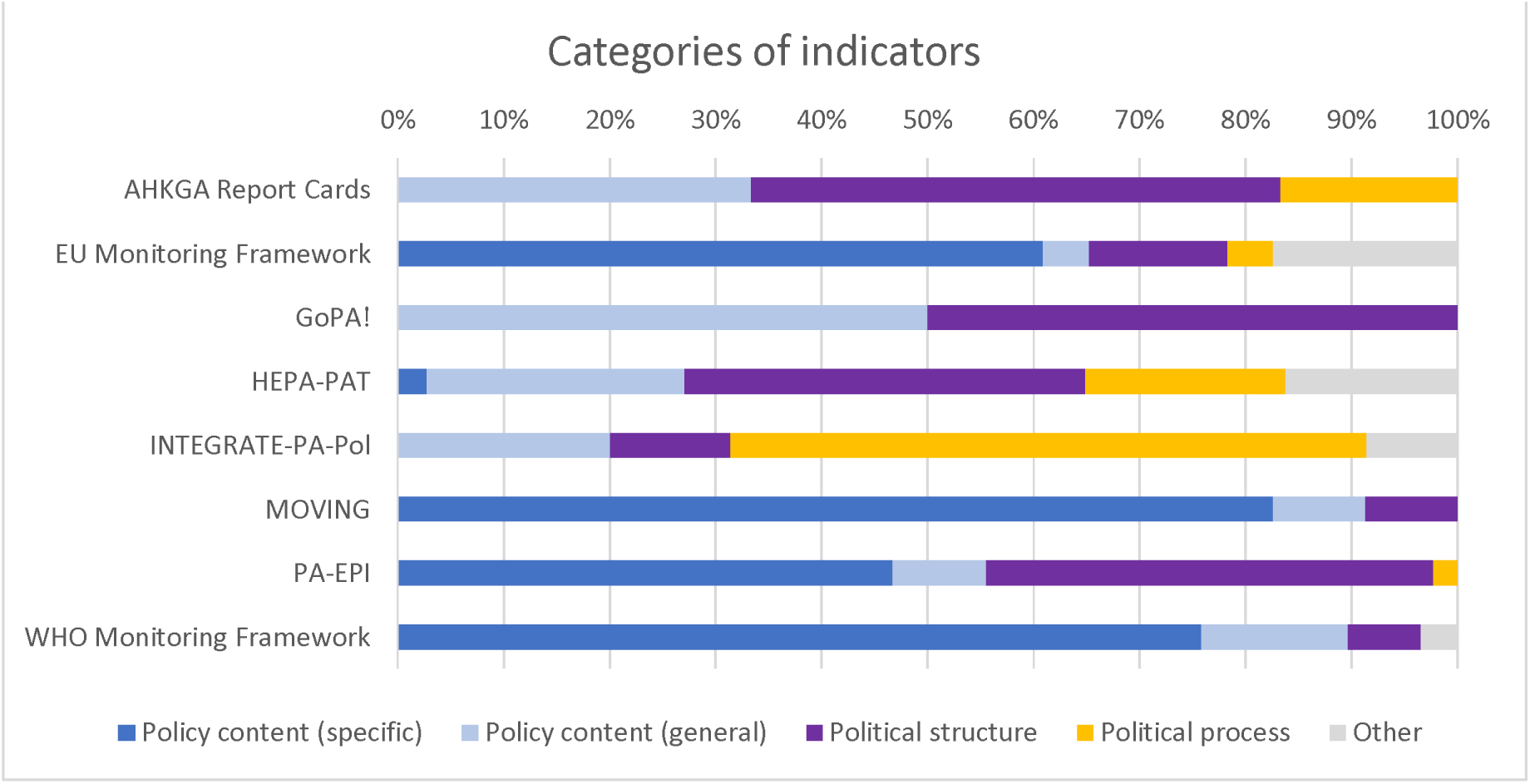
Comparison of indicator categories across tools.

Within these categories, the most frequently identified indicators were:

- *Policy content (specific):* Active travel (n=18), school (n=11), public education/mass media (n=10)
- *Policy content (general):* Key policy documents (n=18), guidelines (n=7)
- *Political structure:* Funding (n=14), surveillance and monitoring system (n=11)
- *Political process:* Policy evaluation (n=9), policy formulation (n=8), policy implementation (n=6)

Most tools that have a strong focus on specific aspects of the policy content address different sectors at a similar level of detail (EU Monitoring Framework, MOVING, PA-EPI). In contrast, the WHO Monitoring Framework emphasizes active travel, which is the primary focus of 11 of its 22 sector-specific indicators (appendix 3).

### Types of indicators

Out of the 202 indicators, 46 (20.3%) audit policies, e.g., ask whether certain policies exist. The content of policies is assessed by 59 indicators (26.0%) and policy implementation by 60 indicators (26.4%). Policy impact is not assessed by any indicator. Thirty-seven indicators were classified as ‘other’.

Figure 2 shows the differences across tools. For three tools, all indicators were categorised into the same type, either content assessment (GoPA!, MOVING) or implementation assessment (PA-EPI). The other five tools combine different types of indicators: AHKGA Report Cards (16.7% audit, 83.3% content assessment); EU Monitoring Framework (43.5% audit, 34.8% content assessment, 8.7% implementation assessment); HEPA-PAT (40.5% audit, 18.9% content assessment, 8.1% implementation assessment); INTEGRATE-PA-Pol (11.4% audit, 11.4% content assessment, 14.3% implementation assessment); and the WHO Monitoring Framework (55.2% audit, 27.6% content assessment, 17.2% implementation assessment).

**Figure 2:**
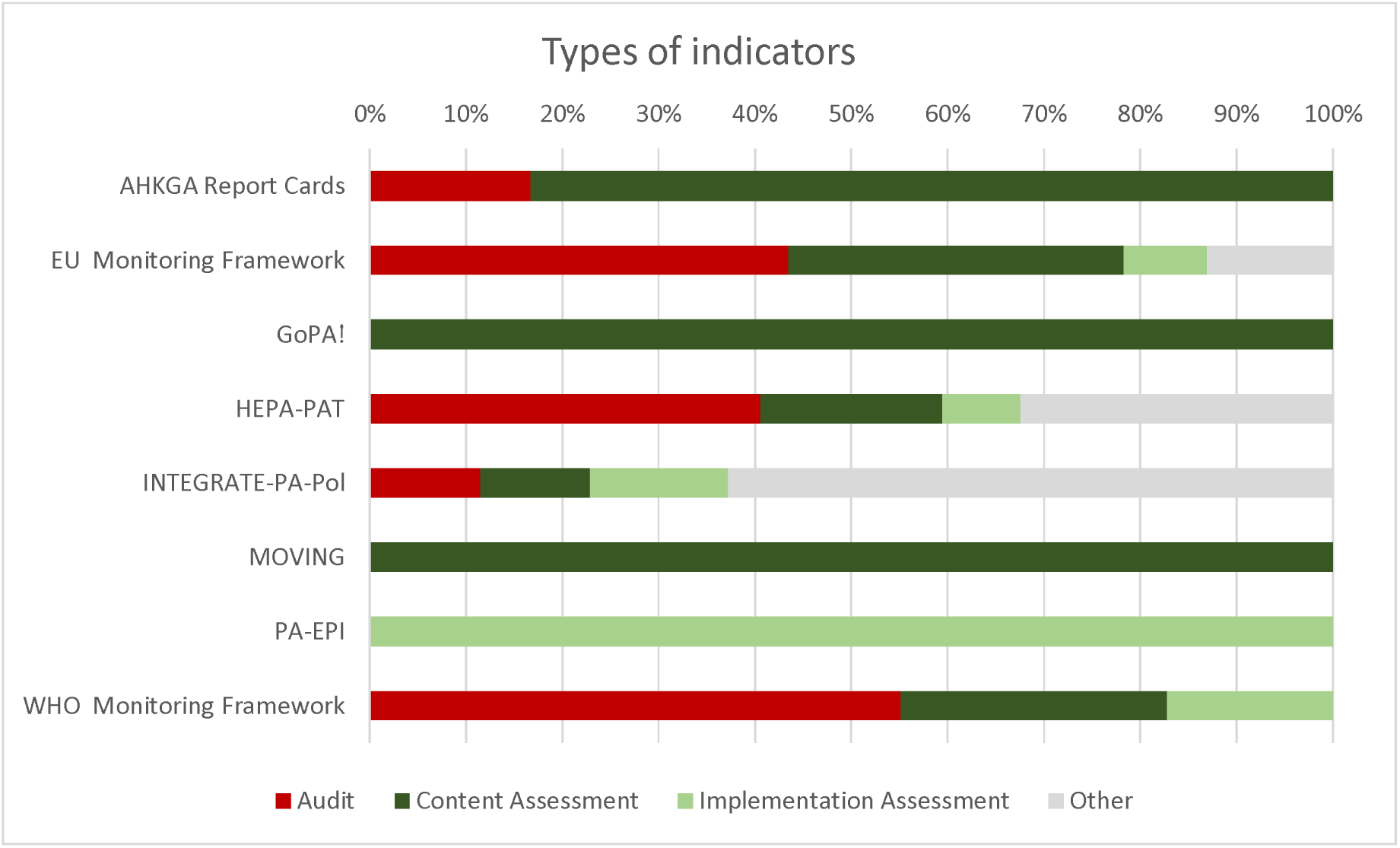
Comparison of indicator types across tools.

## Discussion

This study compared the findings of physical activity policy evaluations across the 27 EU member states. The mean difference in country rank between the highest and lowest was comparatively high (13.0), indicating distinct approaches to physical activity policy evaluation. Mapping 202 indicators showed that each tool focuses on different aspects of physical activity policy, with data collected on policy content (47.6%), the political structure (21.6%), and the political process (13.7%). The study also showed that the tools focus on auditing policies (20.3%), assessing their content (26.0%), and assessing policy implementation (26.4%); none of the indicators assessed policy impact.

One of the key questions raised by this study is how the inconsistencies of country rankings can be explained. It is intuitive to expect that tools analysing the same phenomenon – physical activity policy – would generate similar results and validate each other’s findings. This is true for some countries which have similar ranks across the different tools. For example, Finland and Ireland seem to have comparatively strong physical activity policies, while Cyprus consistently shows room for improvement. For other countries, however, the situation is less clear. Is Spain’s physical activity policy one of the best in Europe (as suggested by the WHO Monitoring Framework) or below average (as indicated by the results of the EU Monitoring Framework)? Similarly, the physical activity policy in Hungary is assessed as comparatively strong by the EU Monitoring Framework and as comparatively weak by the WHO Monitoring Framework. And these are not the only inconsistencies, as shown by the mean difference between the highest and lowest country ranks. This problem is not unique to evaluations of physical activity policies: A previous study comparing the prevalence rates of physical activity across different measurement instruments identified “substantial inconsistencies across and within included physical activity surveillance initiatives, resulting in initiatives contradicting each other” (Aubert et al., 2021). Such contradictions are not helpful for physical activity policy research and advocacy, so it is important to understand the reasons behind these differences. One possible explanation is the varying extent of government involvement across the different tools (Messing et al., 2023). While the EU and WHO Monitoring Frameworks collect data from government representatives of EU member states, the other included tools can be classified as research-driven (Messing et al., 2023). Although researchers may be more critical than government representatives when evaluating public policies, this study’s findings did not confirm that this aspect plays a major role in explaining differences in country rankings: Country rankings were most consistent when a government-driven tool (EU or WHO Monitoring Framework) was compared to a research-driven tool (MOVING).

However, it needs to be considered that these tools have been developed for different purposes, as highlighted by previous research (Klepac Pogrmilovic, O’Sullivan, et al., 2019). This study showed that different physical activity policy evaluation tools collect data on very different indicators, which may help to explain the inconsistencies of country rankings. While some tools collect specific data on policies in particular sectors and settings (e.g., EU-Monitoring Framework, MOVING, PA-EPI, WHO Monitoring Framework), others have a more general focus on available policies. Data on the political structure are mainly collected by tools such as the HEPA-PAT and the PA-EPI, while INTEGRATE-PA-Pol has a unique focus on the political process. From a scientific perspective, this diversity of indicators might be beneficial, as taking different perspectives into account could generate more knowledge on physical activity policy. However, this diversity may be problematic for physical activity advocacy, as clear objectives and a clear agenda for action are essential for achieving advocacy success (Shilton & Milton, 2024). The different evaluation approaches across tools seem to indicate different perceptions of what is important in physical activity policy, even within a specific sector or setting. This is not surprising, given that the public policy literature states that “assumptions of what constitutes [policy] success take many forms” (McConnell, 2010). While there seems to be a general consensus on the importance of different domains for physical activity promotion based on ISPAH’s Eight Investments (Milton et al., 2021), each tool follows a different logic resulting in different indicators. Ideally, different tools would be used for auditing, content assessment and implementation assessment as long as the same or very similar indicators are used. It seems to be essential to reach consensus on key indicators in order to “develop a set of complementary instruments, with each tool collecting detailed information about a specific component”, as previously suggested (Klepac Pogrmilovic, O’Sullivan, et al., 2019). However, achieving consensus may be difficult due to the different purposes of each tool and diverse perspectives on physical activity policy.

### Recommendations for next steps

The authors recommend that each tool clearly communicates its specific objectives and purpose, enabling users to select the tool most suitable for their area of interest. To minimise confusion, it should be transparent what type of data each tool generates and how indicators are measured, for example whether assessments are based on binary yes-no questions or at a higher level of detail. Even when tools assess similar policy domains, their indicators differ (e.g., with regards to the content of active mobility policies). While the tools have the potential to complement each other, there is a risk that evaluation findings may be perceived as inconsistent. Strengthening policy literacy among researchers, practitioners and policymakers is therefore an important priority, as users need to understand not only what evaluation results mean, but also why a country’s policy is assessed as strong or weak, and what actions could help to improve it. Greater clarity is also needed regarding policy terminology and the operationalisation of broad concepts such as “physical activity policy”. A consistent and unambiguous terminology is particularly important when communicating to policymakers, for whom clear messages are essential (Oliver & Cairney, 2019). Future research might help to further develop a common understanding of key policy actions for physical activity promotion, potentially including more regulatory and fiscal policy instruments as opposed to “soft” policies such as recommendations and guidelines (Gelius et al., 2022). This may be particularly valuable for policymakers who often focus on addressing specific barriers to physical activity. Although a holistic cross-sectoral approach can be useful, comparative analyses of policies or interventions targeting specific population groups or policy areas may be more useful for decision-making, particularly when resources are limited. At the same time, broad country comparisons can be valuable from a policy coordination perspective.

As a second recommendation, we would like to highlight the importance of critically reflecting on the data collection process. While some tools use government-reported data, others rely on independent experts or utilize co-production processes between researchers and government officials (Messing et al., 2023). Data collection processes also vary across, and sometimes even within, different tools, and assessments can be problematic if they lack objective criteria or are based solely on the opinion of a single expert. To better understand the strengths and limitations of each tool, future studies may examine whether the application of tools is repeatable and reproducible when data are collected by different individuals or organisations.

A third recommendation is to explore opportunities for greater alignment and linkage between tools. This would involve identifying areas of overlap, i.e. where tools intend to measure the same aspects of physical activity policy (and whether findings for those indicators are consistent across tools). A complementary set of tools – which has previously been suggested in the scientific literature (Klepac Pogrmilovic, O’Sullivan, et al., 2019) – might use the same or closely aligned indicators. For instance, one tool could audit policies, a second could assess policy content, and a third could assess policy implementation, with each building on findings from the previous tool. This would increase the efficiency of the data collection process and ensure that the findings of different tools speak to each other. In this context, initiatives aiming to build consensus in physical activity policy evaluation and/or to support the assignment of weights to the relative importance of different indicators appear to be particularly relevant; ideally, such efforts would be guided by policymakers’ perspectives to enhance their relevance and uptake in practice.

Our fourth recommendation is to address gaps in policy evaluation. Examples of currently underrepresented perspectives include policies in childcare settings (currently assessed by only one of 202 indicators), legal measures, and measures assessing the impact of policies. This suggests scope for formative research to better identify these gaps and develop targeted solutions. Considering policymakers’ interest in improving specific policies or interventions, tools that better incorporate this perspective may be of particular value.

### Strengths and limitations

To the best of our knowledge, this is the first study to compare the findings of physical activity policy evaluation tools. We identified inconsistencies in country rankings and systematically mapped indicators across tools, which may contribute to a better understanding of their differences. As the authors of this study were involved in the data collection or validation processes of most of the analysed tools, this should have prevented any bias for or against any of the included tools. Mapping the indicators also included tools with limited data availability for EU member states, thereby increasing the relevance of this comparative study for future physical activity policy research.

One limitation of this study is that the selection of tools was based on existing reviews and was restricted to tools that had been used in multiple countries. There may be additional approaches to physical activity policy evaluation at the national level. Focusing on national-level tools also excludes the subnational level, even though local and regional levels often have independent roles in promoting physical activity. Another limitation is that not all tools were designed to rank countries, and calculating aggregate scores assumes that each indicator has the same weight. Weighting the indicators based on their importance for physical activity policy, though difficult to determine, may have produced different rankings. Another limitation is that the comparison of country rankings was limited to EU member states, and it is unclear whether similar inconsistencies would have been observed in other world regions. It is also important to note that the intercoder-reliability for the mapping of indicators was significantly higher for the categories (e.g., content, structure, process), indicating that the differentiation by types (e.g., audit, content assessment, implementation assessment) is more subjective and exploratory. Another limitation is that the mapping of indicators simplifies the complex process of policy evaluation, as some indicators addressed more than one category or data type. Some tools such as MOVING or the PA-EPI require a detailed policy audit for each country before an assessment can take place. Another difference is that some tools assessed policy at a single point in time (e.g., MOVING), while others are designed for policy monitoring over time (e.g., AHKGA Report Cards, EU Monitoring Framework, GoPA!). Regardless of these limitations and different purposes, we believe that the ranking of countries and mapping of indicators can help to improve our understanding of the similarities and differences across tools.

### Conclusions

By comparing country rankings and mapping indicators across different physical activity policy evaluation tools, this study identified inconsistencies that have important implications for the users of these tools. Comparing different categories and types of indicators helped to better understand why some countries rank highly in one tool and poorly in another.

Recommendations for future physical activity policy evaluations include clearly communicating the specific purpose of each tool, assessing the potential for linkages between tools, addressing gaps, and reflecting on the reproducibility of the data collection process for each tool.

## Supporting information

Appendix 1: Tool selection process

Appendix 2: Country ranking

Appendix 3: Indicator mapping

## Data Availability

All data produced in the present work are contained in the manuscript.

## Acknowledgements

We would like to acknowledge the consistent support of Benny Cullen (Sport Ireland) and Anne-Laure Caille-Brillet (Federal Ministry of Health, Germany) in the project’s Scientific Advisory Board. In addition, we would like to acknowledge the feedback received from members of the International Public Policy Association on earlier versions of this paper: Emily St. Denny (University of Copenhagen); Philippe Zittoun (University of Lyon); Thilo Bodenstein (Central European University); Achim Kemmerling (University of Erfurt).

## Key points

- Across the 27 EU member states, physical activity policy tools show substantial inconsistencies in country rankings, reflecting distinct approaches to policy evaluation.
- Indicators used in international evaluation tools assess policy content and policy implementation, while neglecting policy impact.
- The diversity of evaluation approaches may advance scientific understanding but can create challenges for clear communication and effective advocacy for physical activity policy.
- For public health policy and practice, establishing a common framework with harmonised indicators is essential to guide more coherent evaluations.

## Appendices

Appendix 1: Tool selection process

Appendix 2: Country ranking

Appendix 3: Indicator mapping

