## Appendix 1: Tool selection process for "Comparing physical activity policy evaluations across 27 EU member states: Diverse indicators, inconsistent findings"

Appendix 1: Selection of tools

*Figure A1.1: Flowchart of tool selection process*

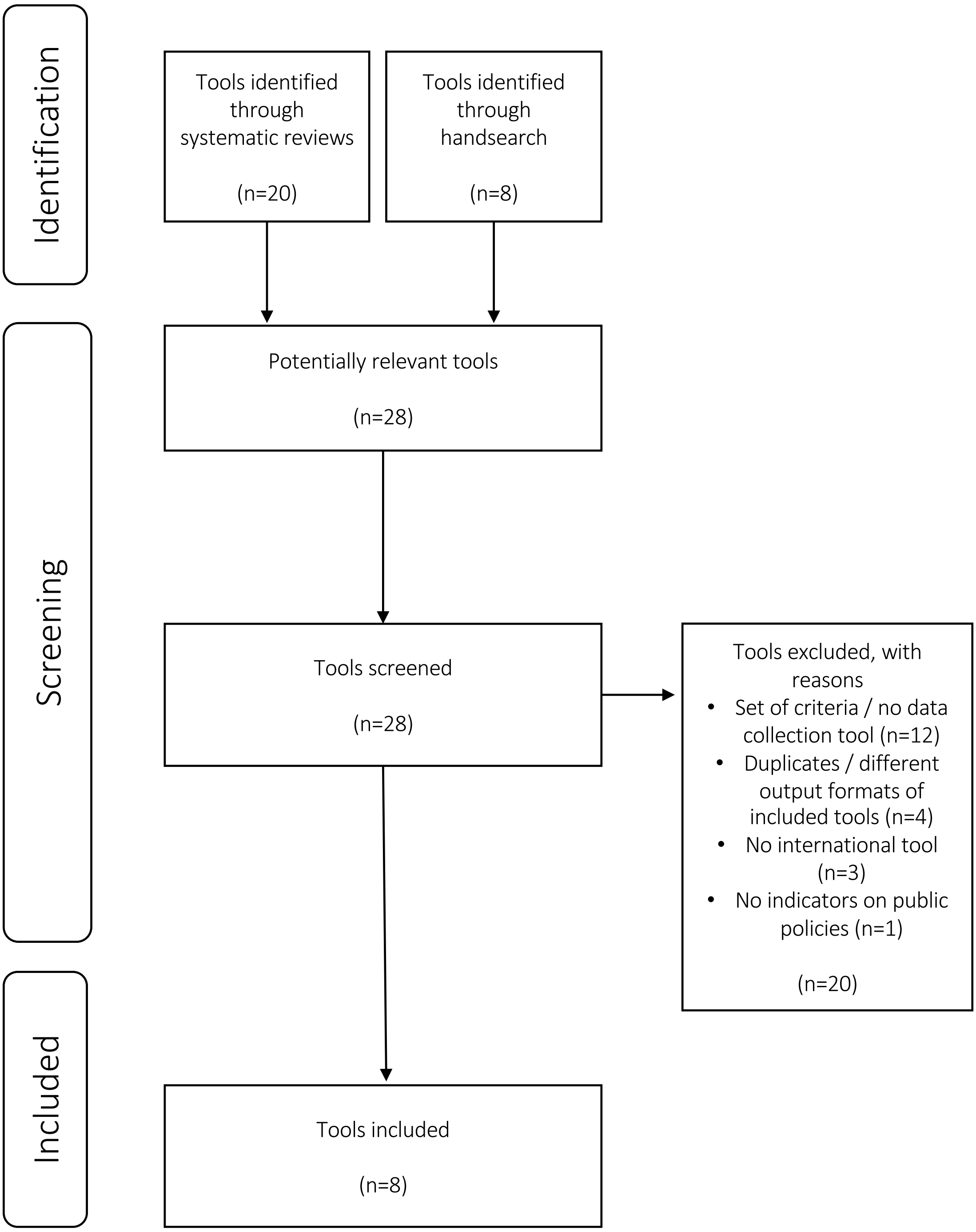

*Table A1.1: List of excluded tools*

| No. | Name of the tool | Author(s) and publication |
|---|---|---|
| 9 | Eurostat | Different online sources (e.g., Eurostat, 2022) |
| 10 | WHO NCD progress monitor report | (WHO, 2025) |
| 11 | Global Obesity Observatory | Website (https://data.worldobesity.org ) |
| 12 | WHO NCD Country Capacity Survey | (WHO, 2026) |
| 13 | GoPE! | (Martins et al., 2025) |
| 14 | A comprehensive PA policy framework | (Shephard et al., 2004) |
| 15 | Analysis Grid for Environments Linked to Obesity (ANGELO) Framework | (Swinburn et al., 1999) |
| 16 | Criteria for successful policy and action plans on physical activity | (Bull, Bauman, et al., 2004; Bull, Bellew, et al., 2004; Schöppe et al., 2004) |
| 17 | Eight aspects identified as being relevant for effective physical activity policies | (Daugbjerg et al., 2009) |
| 18 | Elements of national policy documents | (Branca et al., 2007) |
| 19 | Four cornerstones of a successful national policy framework | (Chalkley & Milton, 2021) |
| 20 | HARDWIRED criteria for successful national physical activity policy | (Bellew et al., 2008) |
| 21 | Important elements of successful physical activity policies and plans | (WHO, 2007a) |
| 22 | Key principles that should guide member states in the development of national physical activity strategies | (WHO, 2007b) |
| 23 | Policy principles for the promotion of healthy diets and physical activity | (WHO, 2003) |
| 24 | Categories for the content analysis of policies | (Christiansen et al., 2014; WHO, 2011) |
| 25 | Analysis of Determinants of Policy Impact (ADEPT) Model | (Rütten et al., 2011) |
| 26 | A graphical, computer-based decision-support tool to help decision makers evaluate policy options relating to PA | (Yancey et al., 2010) |
| 27 | TARGET:PA | (Gelius et al., 2025) |
| 28 | Australian Systems Approach to Physical Activity (ASAPA) tool | (Nau et al., 2019) |
