## Appendix 2: Country ranking for "Comparing physical activity policy evaluations across 27 EU member states: Diverse indicators, inconsistent findings"

### Appendix 2: Country ranking comparison

*Table A2.1: Data sources and data availability of included tools*

| No. | Name of the tool | Data source(s) | No. of EU member states with available data |
|---|---|---|---|
| 1 | AHKGA Report Cards | (Aubert et al., 2022) | 16 |
| 2 | EU Monitoring Framework | (Whiting et al., 2021) | 27 |
| 3 | GoPA! | (Ramirez Varela et al., 2025) | 26 |
| 4 | HEPA-PAT | (Bull et al., 2015; Gelius et al., 2021; Kahlmeier, 2024; Van Hoye et al., 2019) | 11 |
| 5 | INTEGRATE-PA-Pol | (Pratt et al., 2025) | 1 |
| 6 | MOVING | (WCRF & NIPH, 2024) | 25 |
| 7 | PA-EPI | (Heuvelman et al., 2025; Volf et al., 2023) | 2 |
| 8 | WHO Monitoring Framework | (WHO, 2022) | 27 |

*Table A2.2: Calculation of aggregate scores*

| No. | Name of the tool | Description of score calculation | Maximum score |
|---|---|---|---|
| 1 | AHKGA Report Cards | The AHKGA Report Cards assign grades from A+ (best) to F (worst) for the indicator "government". The grades were extracted from the Global Matrix 4.0 (Aubert et al., 2022), and translated into a corresponding number for analysis using the grading rubric of the Active Healthy Kids Global Alliance<br>• Grade A+ \| 15 points<br>• Grade A \| 14 points<br>• Grade A- \| 13 points<br>• Grade B+ \| 12 points<br>• Grade B \| 11 points<br>• Grade B- \| 10 points<br>• Grade C+ \| 9 points<br>• Grade C \| 8 points<br>• Grade C- \| 7 points<br>• Grade D+ \| 6 points<br>• Grade D \| 5 points<br>• Grade D- \| 4 points<br>• Grade F \| 2 points<br>• INC \| Missing value | 15 points |
| 2 | EU Monitoring Framework | The EU Monitoring Framework includes 23 indicators on the implementation of the Council Recommendations on Health Enhancing Physical Activity. Similar to a previous publication (Whiting et al., 2021), a sum score was calculated for the total number of indicators implemented per country:<br>• Indicator implemented \| 1 point<br>• Indicator not implemented \| 0 points | 23 points |

| | | | |
|---|---|---|---|
| 3 | GoPA! | The GoPA! country cards include a physical activity promotion capacity pyramid with data on “policy availability” and “policy implementation”. In each of these two categories, countries are classified as “high”, “medium” or “low”. Data were extracted from the 3rd Physical Activity Almanac (Ramirez Varela et al., 2025), and translated into points:<br>• High \| 3 points<br>• Medium \| 2 points<br>• Low \| 1 point | 6 points |
| 6 | MOVING | The MOVING policy index includes data for the six elements of the MOVING framework which are linked to the following areas: schools (M), workplaces (O), structures (V), transport infrastructure (I), public communication (N) and the healthcare settings (G). The data were extracted from a report of the CO-CREATE project (WCRF & NIPH, 2024). For each element of the MOVING index, countries received a score that was translated into one of five categories:<br>• Excellent (dark green) \| 5 points<br>• Good (light green) \| 4 points<br>• Moderate (yellow) \| 3 points<br>• Fair (orange) \| 2 points<br>• Poor (red) \| 1 point<br>• No policies identified (grey) \| 0 points | 30 points |
| 8 | WHO Monitoring Framework | The WHO Physical Activity Country Profiles – the output of the WHO Monitoring Framework – include 38 indicators in four categories: Active societies (5 indicators), active environments (10 indicators), active people (12 indicators), and active systems (11 indicators). Data were extracted from the Country Profiles of the WHO Global Status Report on Physical Activity (WHO, 2022). For most indicators, response options were “yes” or no”. Some indicators included additional response options. Responses were translated into points as follows:<br>• “Yes and best practice” \| 3 points<br>• “Yes” or “Yes and operational” \| 2 points<br>• “Yes, but not operational” \| 1 point<br>• “No” or “Not available” \| 0 points | 82 points |

*Table A2.3: Aggregate scores across all tools*

| Country | AHKGA Report Card (2022) | EU Monitoring Framework (2024) | GoPA! (2025) | MOVING (2023) | WHO Monitoring Framework (2022) |
|---|---|---|---|---|---|
| Austria | No data | 19 | 5 | 14 | 65 |
| Belgium | No data | 21 | 6 | 16 | 53 |
| Bulgaria | No data | 18 | 6 | 7 | 54 |
| Croatia | 6 | 20 | 6 | 8 | 61 |
| Cyprus | No data | 16 | 3 | No data | 41 |
| Czechia | 6 | 18 | 6 | 8 | 52 |
| Denmark | 12 | 20 | 5 | 17 | 71 |
| Estonia | 11 | 19 | 4 | 11 | 63 |
| Finland | 13 | 23 | 6 | 17 | 67 |
| France | 11 | 22 | 6 | 16 | 71 |
| Germany | INC | 22 | 6 | 17 | 68 |
| Greece | No data | 19 | 3 | 13 | 49 |
| Hungary | 11 | 21 | 6 | 12 | 42 |
| Ireland | 11 | 21 | 6 | 18 | 68 |
| Italy | No data | 17 | 4 | 10 | 62 |
| Latvia | No data | 16 | 3 | 7 | 61 |
| Lithuania | 9 | 19 | 5 | 16 | 71 |
| Luxembourg | No data | 17 | 5 | No data | 66 |
| Malta | No data | 16 | 5 | 6 | 45 |
| Netherlands | No data | 20 | 6 | 13 | 54 |
| Poland | 8 | 19 | 5 | 10 | 66 |
| Portugal | 11 | 20 | 6 | 16 | 67 |
| Romania | No data | 12 | 5 | 4 | 45 |
| Slovakia | 10 | 21 | 5 | 14 | 50 |
| Slovenia | 5 | 17 | 6 | 13 | 65 |
| Spain | 8 | 15 | 6 | 15 | 69 |
| Sweden | 11 | 20 | 5 | 11 | 60 |

*Table A2.3: Percentage of points (in comparison to maximum score of the respective tool)*

| Country | AHKGA Report Card | EU Monitoring Framework | GoPA! | MOVING | WHO Monitoring Framework |
|---|---|---|---|---|---|
| Austria | No data | 83% | 83% | 47% | 79% |
| Belgium | No data | 91% | 100% | 53% | 65% |
| Bulgaria | No data | 78% | 100% | 23% | 66% |
| Croatia | 40% | 87% | 100% | 27% | 74% |
| Cyprus | No data | 70% | 50% | No data | 50% |
| Czechia | 40% | 78% | 100% | 27% | 63% |
| Denmark | 80% | 87% | 83% | 57% | 87% |
| Estonia | 73% | 83% | 67% | 37% | 77% |
| Finland | 87% | 100% | 100% | 57% | 82% |
| France | 73% | 96% | 100% | 53% | 87% |
| Germany | INC | 96% | 100% | 57% | 83% |
| Greece | No data | 83% | 50% | 43% | 60% |
| Hungary | 73% | 91% | 100% | 40% | 51% |
| Ireland | 73% | 91% | 100% | 60% | 83% |
| Italy | No data | 74% | 67% | 33% | 76% |
| Latvia | No data | 70% | 50% | 23% | 74% |
| Lithuania | 60% | 83% | 83% | 53% | 87% |
| Luxembourg | No data | 74% | 83% | No data | 80% |
| Malta | No data | 70% | 83% | 20% | 55% |
| Netherlands | No data | 87% | 100% | 43% | 66% |
| Poland | 53% | 83% | 83% | 33% | 80% |
| Portugal | 73% | 87% | 100% | 53% | 82% |
| Romania | No data | 52% | 83% | 13% | 55% |
| Slovakia | 67% | 91% | 83% | 47% | 61% |
| Slovenia | 33% | 74% | 100% | 43% | 79% |
| Spain | 53% | 65% | 100% | 50% | 84% |
| Sweden | 73% | 87% | 83% | 37% | 73% |
| Average (mean) | 63,6% | 81,8% | 86,4% | 41,2% | 72,5% |

*Table A2.4: Difference between highest and lowest rank*

| Country | Across all five tools | EU, MOVING, WHO | EU, MOVING | EU, WHO | MOVING, WHO |
|---|---|---|---|---|---|
| Austria | 4 | 3 | 3 | 2 | 1 |
| Belgium | 19 | 16 | 1 | 16 | 15 |
| Bulgaria | 21 | 4 | 4 | 0 | 4 |
| Croatia | 19 | 12 | 12 | 7 | 5 |
| Cyprus | 4 | 4 | N/A | 4 | N/A |
| Czechia | 20 | 3 | 2 | 3 | 1 |
| Denmark | 13 | 7 | 6 | 7 | 1 |
| Estonia | 20 | 3 | 3 | 0 | 3 |
| Finland | 6 | 6 | 1 | 6 | 5 |
| France | 4 | 4 | 3 | 1 | 4 |
| Germany | 4 | 3 | 0 | 3 | 3 |
| Greece | 13 | 11 | 1 | 10 | 11 |
| Hungary | 25 | 22 | 11 | 22 | 11 |
| Ireland | 4 | 4 | 3 | 1 | 4 |
| Italy | 9 | 6 | 2 | 6 | 4 |
| Latvia | 10 | 8 | 1 | 8 | 7 |
| Lithuania | 13 | 12 | 8 | 12 | 4 |
| Luxembourg | 11 | 11 | N/A | 11 | N/A |
| Malta | 10 | 1 | 1 | 1 | 0 |
| Netherlands | 17 | 10 | 4 | 10 | 6 |
| Poland | 9 | 9 | 5 | 4 | 9 |
| Portugal | 7 | 3 | 3 | 1 | 2 |
| Romania | 13 | 3 | 2 | 3 | 1 |
| Slovakia | 18 | 18 | 6 | 18 | 12 |
| Slovenia | 19 | 9 | 8 | 9 | 1 |
| Spain | 25 | 22 | 17 | 22 | 5 |
| Sweden | 14 | 9 | 8 | 9 | 1 |
| Average (mean) | 13,0 | 8,3 | 4,6 | 7,3 | 4,8 |
