## Appendix 3: Indicator mapping for "Comparing physical activity policy evaluations across 27 EU member states: Diverse indicators, inconsistent findings"

*Table A3.1: Coding tree for indicator categories*

| Category (Level 1) | Category (Level 2) | Sub-Category (Level 3) |
|---|---|---|
| Policy content | Specific (sector/setting) | Active travel |
| | | Active urban design |
| | | Childcare |
| | | Community |
| | | Healthcare |
| | | Public education / mass media |
| | | School |
| | | Sports for all |
| | | Workplace |
| | General | Key policy documents |
| | | Guidelines |
| | | Comprehensiveness |
| | | Goals / actions / tasks |
| Political structure | N/A | Coordination mechanism |
| | | Funding |
| | | Government structure |
| | | Health in all policies |
| | | Leadership |
| | | Surveillance and monitoring system |
| | | Workforce development |
| Political process | N/A | Agenda setting |
| | | Policy formulation |
| | | Decision making / adoption |
| | | Policy implementation |
| | | Policy evaluation |
| Other | N/A | Behavioural indicator |
| | | Other indicator |

*Table A3.2: Coding tree for indicator types*

| Type (Level 1) | Description |
|---|---|
| Audit | Auditing is an inquiry about a certain aspect of policy but not rating, grading, judging, or evaluating it. An example of a questionnaire item used for this purpose is: „Does Australia have a national PA strategy?" (Klepac Pogrmilovic et al., 2019). |
| Content assessment | Content assessment or evaluation may focus on a number of different aspects of policy content, e.g. the core components and implementation requirements of the policy or the evidence base supporting the policy (CDC, 2012). |
| Implementation assessment | Implementation assessment or evaluation may focus on a number of different areas including components of logic models – such as inputs, activities and outputs – or facilitators of and barriers to implementation (CDC, 2012). |
| Impact assessment | Impact assessment of evaluation may be on short-term, intermediate, and long-term outcomes and impacts, costs of implementing the policy, or cost savings resulting from policy implementation (CDC, 2012). |

*Table A3.3: Categories of indicators (frequency)*

| | AHKGA Report Cards | EU | GoPA! | HEPA-PAT | INTEGRATE | MOVING | PA-EPI | WHO | Total |
|---|---|---|---|---|---|---|---|---|---|
| **Policy content (specific)** | | **14** | | **1** | | **19** | **21** | **22** | **77** |
| Active travel | | | | | | 4 | 3 | 11 | 18 |
| Active urban design | | 1 | | | | 5 | 3 | | 9 |
| Childcare | | | | | | | | 1 | 1 |
| Community | | 1 | | | | 1 | 2 | 3 | 7 |
| Healthcare | | 2 | | | | 3 | 2 | 2 | 9 |
| Public education / mass media | | 1 | | 1 | | 2 | 2 | 4 | 10 |
| School | | 4 | | | | 3 | 4 | | 11 |
| Sport for all | | 3 | | | | | 3 | | 6 |
| Workplace | | 2 | | | | 1 | 2 | 1 | 6 |
| **Policy content (general)** | **2** | **1** | **2** | **9** | **7** | **2** | **4** | **4** | **31** |
| Comprehensiveness | | | | 1 | | | | | 1 |
| Goals / actions / tasks | 1 | | 1 | 2 | | | | 1 | 5 |
| Guidelines | | 1 | | 2 | | 1 | 2 | 1 | 7 |
| Key policy documents | 1 | | 1 | 4 | 7 | 1 | 2 | 2 | 18 |
| **Political structure** | **3** | **3** | **2** | **14** | **4** | **2** | **19** | **2** | **49** |
| Coordination mechanism | | 1 | | 3 | | | 3 | 1 | 8 |
| Funding | 1 | 1 | 1 | 2 | 4 | 1 | 4 | | 14 |
| Government structure | | | 1 | 3 | | | 1 | | 5 |
| Health in all policies | | | | | | | 2 | | 2 |
| Leadership | 1 | | | 3 | | | 1 | | 5 |
| Surveillance and monitoring system | 1 | 1 | | 3 | | | 5 | 1 | 11 |
| Workforce development | | | | | | 1 | 3 | | 4 |

| | AHKGA Report Cards | EU | GoPA! | HEPA-PAT | INTEGRATE | MOVING | PA-EPI | WHO | Total |
|---|---|---|---|---|---|---|---|---|---|
| **Political process** | **1** | **1** | | **7** | **21** | | **1** | | **31** |
| Agenda setting | | | | | 4 | | | | 4 |
| Policy formulation | | | | 3 | 4 | | 1 | | 8 |
| Decision making / adoption | | | | | 4 | | | | 4 |
| Policy implementation | | | | 1 | 5 | | | | 6 |
| Policy evaluation | 1 | 1 | | 3 | 4 | | | | 9 |
| **Other** | | **4** | | **6** | **3** | | | **1** | **14** |
| Behavioural indicator | | 3 | | | | | | | 3 |
| Other indicator | | 1 | | 6 | 3 | | | 1 | 11 |
| **Total** | **6** | **23** | **4** | **37** | **35** | **23** | **45** | **29** | **202** |

*Table A3.4: Types of indicators (frequency)*

| | AHKGA Report Cards | EU | GoPA! | HEPA-PAT | INTEGRATE | MOVING | PA-EPI | WHO | Total |
|---|---|---|---|---|---|---|---|---|---|
| Audit | 1 | 10 | | 15 | 4 | | | 16 | 46 |
| Content assessment | 5 | 8 | 4 | 7 | 4 | 23 | | 8 | 59 |
| Implementation assessment | | 2 | | 3 | 5 | | 45 | 5 | 60 |
| Impact assessment | | | | | | | | | 0 |
| Other | | 3 | | 12 | 22 | | | | 37 |
| **Total** | **6** | **23** | **4** | **37** | **35** | **23** | **45** | **29** | **202** |

*Table A3.5: Categories and types of indicators (list)*

| No. | Tool | Category (Level 1) | Category (Level 2) | Sub-Category (Level 3) | Type |
|---|---|---|---|---|---|
| 1 | AHKGA | Policy content | Policy documents | Key policy documents | Audit |
| 2 | AHKGA | Policy content | Policy documents | Goals / actions / tasks | Content assessment |
| 3 | AHKGA | Political structure | Political structure | Leadership | Content assessment |
| 4 | AHKGA | Political structure | Political structure | Surveillance and monitoring system | Content assessment |
| 5 | AHKGA | Political structure | Political structure | Funding | Content assessment |
| 6 | AHKGA | Political process | Political process | Policy evaluation | Content assessment |
| 7 | EU | Policy content | Policy documents | Guidelines | Audit |
| 8 | EU | Other | Other | Behavioural indicator | Other |
| 9 | EU | Other | Other | Behavioural indicator | Other |
| 10 | EU | Political structure | Political structure | Coordination mechanism | Content assessment |
| 11 | EU | Political structure | Political structure | Funding | Implementation assessment |
| 12 | EU | Policy content | Sector/setting | Sport for all | Audit |
| 13 | EU | Policy content | Sector/setting | Sport for all | Audit |
| 14 | EU | Policy content | Sector/setting | Sport for all | Audit |
| 15 | EU | Other | Other | Other indicator | Content assessment |
| 16 | EU | Political structure | Political structure | Surveillance and monitoring system | Content assessment |
| 17 | EU | Policy content | Sector/setting | Healthcare | Audit |
| 18 | EU | Policy content | Sector/setting | Healthcare | Content assessment |
| 19 | EU | Policy content | Sector/setting | School | Content assessment |
| 20 | EU | Policy content | Sector/setting | School | Audit |
| 21 | EU | Policy content | Sector/setting | School | Content assessment |
| 22 | EU | Policy content | Sector/setting | School | Content assessment |
| 23 | EU | Other | Other | Behavioural indicator | Other |
| 24 | EU | Policy content | Sector/setting | Active urban design | Implementation assessment |

| No. | Tool | Category (Level 1) | Category (Level 2) | Sub-Category (Level 3) | Type |
|---|---|---|---|---|---|
| 25 | EU | Policy content | Sector/setting | Workplace | Audit |
| 26 | EU | Policy content | Sector/setting | Workplace | Audit |
| 27 | EU | Policy content | Sector/setting | Community | Audit |
| 28 | EU | Political process | Political process | Policy evaluation | Content assessment |
| 29 | EU | Policy content | Sector/setting | Public education / mass media | Audit |
| 30 | GoPA | Policy content | Policy documents | Goals / actions / tasks | Content assessment |
| 31 | GoPA | Political structure | Political structure | Funding | Content assessment |
| 32 | GoPA | Policy content | Policy documents | Key policy documents | Content assessment |
| 33 | GoPA | Political structure | Political structure | Government structure | Content assessment |
| 34 | HEPA-PAT | Political structure | Political structure | Government structure | Other |
| 35 | HEPA-PAT | Political structure | Political structure | Government structure | Other |
| 36 | HEPA-PAT | Political structure | Political structure | Government structure | Audit |
| 37 | HEPA-PAT | Other | Other | Other indicator | Audit |
| 38 | HEPA-PAT | Political structure | Political structure | Leadership | Audit |
| 39 | HEPA-PAT | Political structure | Political structure | Leadership | Audit |
| 40 | HEPA-PAT | Political structure | Political structure | Coordination mechanism | Audit |
| 41 | HEPA-PAT | Political structure | Political structure | Coordination mechanism | Audit |
| 42 | HEPA-PAT | Policy content | Policy documents | Key policy documents | Audit |
| 43 | HEPA-PAT | Policy content | Policy documents | Key policy documents | Content assessment |
| 44 | HEPA-PAT | Political process | Political process | Policy formulation | Other |
| 45 | HEPA-PAT | Policy content | Policy documents | Comprehensiveness | Content assessment |
| 46 | HEPA-PAT | Political process | Political process | Policy formulation | Other |
| 47 | HEPA-PAT | Political process | Political process | Policy formulation | Other |
| 48 | HEPA-PAT | Policy content | Policy documents | Key policy documents | Audit |
| 49 | HEPA-PAT | Policy content | Policy documents | Key policy documents | Content assessment |

| No. | Tool | Category (Level 1) | Category (Level 2) | Sub-Category (Level 3) | Type |
|---|---|---|---|---|---|
| 50 | HEPA-PAT | Other | Other | Other indicator | Content assessment |
| 51 | HEPA-PAT | Policy content | Sector/setting | Public education / mass media | Audit |
| 52 | HEPA-PAT | Political process | Political process | Policy implementation | Implementation assessment |
| 53 | HEPA-PAT | Policy content | Policy documents | Guidelines | Audit |
| 54 | HEPA-PAT | Policy content | Policy documents | Guidelines | Audit |
| 55 | HEPA-PAT | Policy content | Policy documents | Goals | Content assessment |
| 56 | HEPA-PAT | Policy content | Policy documents | Goals | Content assessment |
| 57 | HEPA-PAT | Political structure | Political structure | Surveillance and monitoring system | Content assessment |
| 58 | HEPA-PAT | Political structure | Political structure | Surveillance and monitoring system | Other |
| 59 | HEPA-PAT | Political structure | Political structure | Surveillance and monitoring system | Other |
| 60 | HEPA-PAT | Political process | Political process | Policy evaluation | Audit |
| 61 | HEPA-PAT | Political process | Political process | Policy evaluation | Audit |
| 62 | HEPA-PAT | Political process | Political process | Policy evaluation | Audit |
| 63 | HEPA-PAT | Political structure | Political structure | Funding | Implementation assessment |
| 64 | HEPA-PAT | Political structure | Political structure | Funding | Implementation assessment |
| 65 | HEPA-PAT | Political structure | Political structure | Leadership | Other |
| 66 | HEPA-PAT | Political structure | Political structure | Coordination mechanism | Audit |
| 67 | HEPA-PAT | Other | Other | Other indicator | Other |
| 68 | HEPA-PAT | Other | Other | Other indicator | Other |
| 69 | HEPA-PAT | Other | Other | Other indicator | Other |
| 70 | HEPA-PAT | Other | Other | Other indicator | Other |
| 71 | INTEGRATE | Policy content | Policy documents | Key policy documents | Audit |
| 72 | INTEGRATE | Policy content | Policy documents | Key policy documents | Audit |
| 73 | INTEGRATE | Policy content | Policy documents | Key policy documents | Audit |
| 74 | INTEGRATE | Policy content | Policy documents | Key policy documents | Audit |

| No. | Tool | Category (Level 1) | Category (Level 2) | Sub-Category (Level 3) | Type |
|---|---|---|---|---|---|
| 75 | INTEGRATE | Policy content | Policy documents | Key policy documents | Content assessment |
| 76 | INTEGRATE | Political process | Political process | Agenda setting | Other |
| 77 | INTEGRATE | Political process | Political process | Policy formulation | Other |
| 78 | INTEGRATE | Political process | Political process | Decision making / adoption | Other |
| 79 | INTEGRATE | Political process | Political process | Policy implementation | Other |
| 80 | INTEGRATE | Political process | Political process | Policy evaluation | Other |
| 81 | INTEGRATE | Political process | Political process | Agenda setting | Other |
| 82 | INTEGRATE | Political process | Political process | Policy formulation | Other |
| 83 | INTEGRATE | Political process | Political process | Decision making / adoption | Other |
| 84 | INTEGRATE | Political process | Political process | Policy implementation | Other |
| 85 | INTEGRATE | Political process | Political process | Policy evaluation | Other |
| 86 | INTEGRATE | Political process | Political process | Agenda setting | Other |
| 87 | INTEGRATE | Political process | Political process | Policy formulation | Other |
| 88 | INTEGRATE | Political process | Political process | Decision making / adoption | Other |
| 89 | INTEGRATE | Political process | Political process | Policy implementation | Other |
| 90 | INTEGRATE | Political process | Political process | Policy evaluation | Other |
| 91 | INTEGRATE | Other | Other | Other indicator | Other |
| 92 | INTEGRATE | Political process | Political process | Agenda setting | Other |
| 93 | INTEGRATE | Political process | Political process | Policy formulation | Other |
| 94 | INTEGRATE | Political process | Political process | Decision making / adoption | Other |
| 95 | INTEGRATE | Political process | Political process | Policy implementation | Other |
| 96 | INTEGRATE | Political process | Political process | Policy evaluation | Other |
| 97 | INTEGRATE | Other | Other | Other indicator | Other |
| 98 | INTEGRATE | Policy content | Policy documents | Key policy documents | Content assessment |
| 99 | INTEGRATE | Other | Other | Other indicator | Content assessment |

| No. | Tool | Category (Level 1) | Category (Level 2) | Sub-Category (Level 3) | Type |
|---|---|---|---|---|---|
| 100 | INTEGRATE | Policy content | Policy documents | Key policy documents | Content assessment |
| 101 | INTEGRATE | Political process | Political process | Policy implementation | Implementation assessment |
| 102 | INTEGRATE | Political structure | Political structure | Funding | Implementation assessment |
| 103 | INTEGRATE | Political structure | Political structure | Funding | Implementation assessment |
| 104 | INTEGRATE | Political structure | Political structure | Funding | Implementation assessment |
| 105 | INTEGRATE | Political structure | Political structure | Funding | Implementation assessment |
| 106 | MOVING | Policy content | Sector/setting | School | Content assessment |
| 107 | MOVING | Policy content | Sector/setting | School | Content assessment |
| 108 | MOVING | Policy content | Sector/setting | School | Content assessment |
| 109 | MOVING | Policy content | Sector/setting | Community | Content assessment |
| 110 | MOVING | Policy content | Policy documents | Key policy documents | Content assessment |
| 111 | MOVING | Political structure | Political structure | Funding | Content assessment |
| 112 | MOVING | Political structure | Political structure | Workforce development | Content assessment |
| 113 | MOVING | Policy content | Sector/setting | Workplace | Content assessment |
| 114 | MOVING | Policy content | Sector/setting | Active urban design | Content assessment |
| 115 | MOVING | Policy content | Sector/setting | Active urban design | Content assessment |
| 116 | MOVING | Policy content | Sector/setting | Active urban design | Content assessment |
| 117 | MOVING | Policy content | Sector/setting | Active travel | Content assessment |
| 118 | MOVING | Policy content | Sector/setting | Active urban design | Content assessment |
| 119 | MOVING | Policy content | Sector/setting | Active urban design | Content assessment |
| 120 | MOVING | Policy content | Sector/setting | Active travel | Content assessment |
| 121 | MOVING | Policy content | Sector/setting | Active travel | Content assessment |
| 122 | MOVING | Policy content | Sector/setting | Public education / mass media | Content assessment |
| 123 | MOVING | Policy content | Sector/setting | Active travel | Content assessment |
| 124 | MOVING | Policy content | Sector/setting | Public education / mass media | Content assessment |

| No. | Tool | Category (Level 1) | Category (Level 2) | Sub-Category (Level 3) | Type |
|---|---|---|---|---|---|
| 125 | MOVING | Policy content | Policy documents | Guidelines | Content assessment |
| 126 | MOVING | Policy content | Sector/setting | Healthcare | Content assessment |
| 127 | MOVING | Policy content | Sector/setting | Healthcare | Content assessment |
| 128 | MOVING | Policy content | Sector/setting | Healthcare | Content assessment |
| 129 | PA-EPI | Policy content | Sector/setting | School | Implementation assessment |
| 130 | PA-EPI | Policy content | Sector/setting | School | Implementation assessment |
| 131 | PA-EPI | Policy content | Sector/setting | School | Implementation assessment |
| 132 | PA-EPI | Policy content | Sector/setting | School | Implementation assessment |
| 133 | PA-EPI | Policy content | Sector/setting | Active travel | Implementation assessment |
| 134 | PA-EPI | Policy content | Sector/setting | Active travel | Implementation assessment |
| 135 | PA-EPI | Policy content | Sector/setting | Active travel | Implementation assessment |
| 136 | PA-EPI | Policy content | Sector/setting | Active urban design | Implementation assessment |
| 137 | PA-EPI | Policy content | Sector/setting | Active urban design | Implementation assessment |
| 138 | PA-EPI | Policy content | Sector/setting | Active urban design | Implementation assessment |
| 139 | PA-EPI | Policy content | Sector/setting | Healthcare | Implementation assessment |
| 140 | PA-EPI | Policy content | Sector/setting | Healthcare | Implementation assessment |
| 141 | PA-EPI | Policy content | Sector/setting | Public education / mass media | Implementation assessment |
| 142 | PA-EPI | Policy content | Sector/setting | Public education / mass media | Implementation assessment |
| 143 | PA-EPI | Policy content | Sector/setting | Community | Implementation assessment |
| 144 | PA-EPI | Policy content | Sector/setting | Community | Implementation assessment |
| 145 | PA-EPI | Policy content | Sector/setting | Sport for all | Implementation assessment |
| 146 | PA-EPI | Policy content | Sector/setting | Sport for all | Implementation assessment |
| 147 | PA-EPI | Policy content | Sector/setting | Sport for all | Implementation assessment |
| 148 | PA-EPI | Policy content | Sector/setting | Workplace | Implementation assessment |
| 149 | PA-EPI | Policy content | Sector/setting | Workplace | Implementation assessment |

| No. | Tool | Category (Level 1) | Category (Level 2) | Sub-Category (Level 3) | Type |
|---|---|---|---|---|---|
| 150 | PA-EPI | Political structure | Political structure | Leadership | Implementation assessment |
| 151 | PA-EPI | Policy content | Policy documents | Key policy documents | Implementation assessment |
| 152 | PA-EPI | Policy content | Policy documents | Key policy documents | Implementation assessment |
| 153 | PA-EPI | Policy content | Policy documents | Guidelines | Implementation assessment |
| 154 | PA-EPI | Political structure | Political structure | Government structure | Implementation assessment |
| 155 | PA-EPI | Political process | Political process | Policy formulation | Implementation assessment |
| 156 | PA-EPI | Policy content | Policy documents | Guidelines | Implementation assessment |
| 157 | PA-EPI | Political structure | Political structure | Coordination mechanism | Implementation assessment |
| 158 | PA-EPI | Political structure | Political structure | Surveillance and monitoring system | Implementation assessment |
| 159 | PA-EPI | Political structure | Political structure | Surveillance and monitoring system | Implementation assessment |
| 160 | PA-EPI | Political structure | Political structure | Surveillance and monitoring system | Implementation assessment |
| 161 | PA-EPI | Political structure | Political structure | Surveillance and monitoring system | Implementation assessment |
| 162 | PA-EPI | Political structure | Political structure | Surveillance and monitoring system | Implementation assessment |
| 163 | PA-EPI | Political structure | Political structure | Funding | Implementation assessment |
| 164 | PA-EPI | Political structure | Political structure | Funding | Implementation assessment |
| 165 | PA-EPI | Political structure | Political structure | Funding | Implementation assessment |
| 166 | PA-EPI | Political structure | Political structure | Funding | Implementation assessment |
| 167 | PA-EPI | Political structure | Political structure | Coordination mechanism | Implementation assessment |
| 168 | PA-EPI | Political structure | Political structure | Coordination mechanism | Implementation assessment |
| 169 | PA-EPI | Political structure | Political structure | Workforce development | Implementation assessment |
| 170 | PA-EPI | Political structure | Political structure | Workforce development | Implementation assessment |
| 171 | PA-EPI | Political structure | Political structure | Workforce development | Implementation assessment |
| 172 | PA-EPI | Political structure | Political structure | Health in all policies | Implementation assessment |
| 173 | PA-EPI | Political structure | Political structure | Health in all policies | Implementation assessment |
| 174 | WHO | Policy content | Sector/setting | Public education / mass media | Implementation assessment |

| No. | Tool | Category (Level 1) | Category (Level 2) | Sub-Category (Level 3) | Type |
|---|---|---|---|---|---|
| 175 | WHO | Policy content | Sector/setting | Public education / mass media | Implementation assessment |
| 176 | WHO | Policy content | Sector/setting | Public education / mass media | Implementation assessment |
| 177 | WHO | Policy content | Sector/setting | Public education / mass media | Implementation assessment |
| 178 | WHO | Policy content | Sector/setting | Community | Implementation assessment |
| 179 | WHO | Policy content | Sector/setting | Active travel | Audit |
| 180 | WHO | Policy content | Sector/setting | Active travel | Audit |
| 181 | WHO | Policy content | Sector/setting | Active travel | Content assessment |
| 182 | WHO | Policy content | Sector/setting | Active travel | Audit |
| 183 | WHO | Policy content | Sector/setting | Active travel | Content assessment |
| 184 | WHO | Policy content | Sector/setting | Active travel | Content assessment |
| 185 | WHO | Policy content | Sector/setting | Active travel | Content assessment |
| 186 | WHO | Policy content | Sector/setting | Active travel | Content assessment |
| 187 | WHO | Policy content | Sector/setting | Active travel | Content assessment |
| 188 | WHO | Policy content | Sector/setting | Active travel | Content assessment |
| 189 | WHO | Policy content | Sector/setting | Healthcare | Audit |
| 190 | WHO | Policy content | Sector/setting | Childcare | Audit |
| 191 | WHO | Policy content | Sector/setting | Workplace | Audit |
| 192 | WHO | Policy content | Sector/setting | Community | Audit |
| 193 | WHO | Policy content | Sector/setting | Community | Audit |
| 194 | WHO | Policy content | Sector/setting | Active travel | Audit |
| 195 | WHO | Other | Other | Other indicator | Audit |
| 196 | WHO | Policy content | Sector/setting | Healthcare | Audit |
| 197 | WHO | Policy content | Policy documents | Key policy documents | Audit |
| 198 | WHO | Policy content | Policy documents | Key policy documents | Audit |
| 199 | WHO | Policy content | Policy documents | Guidelines | Audit |

| No. | Tool | Category (Level 1) | Category (Level 2) | Sub-Category (Level 3) | Type |
|---|---|---|---|---|---|
| 200 | WHO | Policy content | Policy documents | Goals | Content assessment |
| 201 | WHO | Political structure | Political structure | Coordination mechanism | Audit |
| 202 | WHO | Political structure | Political structure | Surveillance and monitoring system | Audit |

*Table A3.6: Wording of indicators (list)*

| No. | Tool | Indicator / Category | Description |
|---|---|---|---|
| 1 | AHKGA | Number and breadth of relevant policies:<br>Policies/strategies/action plans that reference physical activity | Policy number & policy breadth (no. of sectors) |
| 2 | AHKGA | Identified supporting actions:<br>Strategic documents with specific actions that promote physical activity | No. of policies with identifiable actions |
| 3 | AHKGA | Identified accountable organization(s):<br>Discreet organizations specifically identified as responsible for delivery ofa ctions | Proportion (%) of policies with identified responsibilities for delivery of actions |
| 4 | AHKGA | Identifiable reporting structures:<br>Strategic documents with explicit reporting systems including frequency and format of reports | Proportion (%) of policies with identified systems for reporting delivery of actions |
| 5 | AHKGA | Identified funding:<br>Explicit references to funding to support identified actions | Proportion (%) of policies with identified funding sources |
| 6 | AHKGA | Monitoring and evaluation plan:<br>Explicit reference to monitoring and evaluations of progress and impact of the policy | Proportion (%) of included policies with identified systems for monitoring and evaluation |
| 7 | EU | 1 National recommendation on physical activity for health | Yes/no |
| 8 | EU | 2. Adults reaching the minimum WHO recommendation on physical activity for health | Percentage of adults reaching a minimum of 150 minutes of moderate-intensity physical activity per week, or 75 minutes of vigorous- intensity activity, or an equivalent combination |
| 9 | EU | 3. Children and adolescents reaching the minimum WHO recommendation on physical activity for health | Percentage of children and adolescents reaching at least 60 minutes of mode-rate- to vigorous-intensity physical activity daily or on at least 5 days / week |

| No. | Tool | Indicator / Category | Description |
| --- | --- | --- | --- |
| 10 | EU | 4 National coordination mechanism on HEPA promotion | Yes/no; if yes:<br>- Name? Since when in place?<br>- Which sectors and stakeholders are participating (pre-defined list)<br>- Which is the leading institution?<br>- Has funding been allocated to this coordinating mechanism? If yes: o total funding; o per capita; o by gross domestic product at PPP per capita, in Euros |
| 11 | EU | 5 Funding allocated specifically to HEPA promotion | By sector (health, sport, transport etc.):<br>- total funding;<br>- per capita;<br>- by gross domestic product at PPP per capita, in Euros |
| 12 | EU | 6 National sport for all policy and/or action plan | Yes/no; if yes: name, status, issuing body, policy areas covered, web-link. |
| 13 | EU | 7 Sports Clubs for Health Programme | Implementation of the guidelines developed by HEPA Europe/TAFISA project: yes/no; if yes, description |
| 14 | EU | 8 Framework to support opportunities to increase access to recreational or exercise facilities for low socio-economic groups | Existence of a framework: yes/ foreseen within the next 2 years/no; and if yes, description |
| 15 | EU | 9 Target groups addressed by the national HEPA policy | By target group (groups in particular need of physical activity (e.g. low socio-economic groups, people with low levels of PA, elderly, ethnic minorities etc.) |
| 16 | EU | 10 Monitoring and surveillance of physical activity | Physical activity included in the national health monitoring system: yes/no If yes: name of the survey, year, measured items, age groups, socioeconomics, link to survey |
| 17 | EU | 11 Counselling on physical activity | Counselling on physical activity: yes / no If yes: reimbursed as part of primary health care services: yes/no |
| 18 | EU | 12 Training on physical activity in curriculum for health professionals | '- number of hours for nurses, doctors̈ - mandatory or optional - clear assessment and accreditation<br>structures to reflect the learning outcomes of the subject |
| 19 | EU | 13 Physical education in primary and secondary schools | - number of hours per school level - mandatory or optional - national or sub-national regulation |

| No. | Tool | Indicator / Category | Description |
|---|---|---|---|
| 20 | EU | 14 Schemes for school-related physical activity promotion | Existence of a national or sub-national (where relevant#) scheme Yes/no - active school breaks - active breaks during school lessons - after-school HEPA programmes (at schools, at sport clubs, in communities) |
| 21 | EU | 15 HEPA in training of physical education teachers | HEPA being a module in training of PE teachers at bachelor's and/or master's degree level: yes/no; mandatory/optional |
| 22 | EU | 16 Schemes promoting active travel to school | National or sub-national (where relevant#) schemes to promote active travel to school (e.g. walking buses, cycling): Yes/no, if yes: description |
| 23 | EU | 17 Level of cycling / walking | Main mode of transport used for your daily activities (car, motorbike, public transport, walking, cycling, other) |
| 24 | EU | 18 European Guidelines for improving infrastructures for Leisure-Time Physical Activity | European Guidelines for improving Infrastructures for Leisure-Time Physical Activity (addressing sport infrastructure, leisure-time infrastructure and urban and green spaces) being applied systematically to plan, build and manage infrastructures: Yes / not yet but foreseen within the next 2 years / no |
| 25 | EU | 19 Schemes to promote active travel to work | Existence of a national or sub-national (where relevant#) incentive scheme for companies or employees to promote active travel to work (e.g. walking, cycling): yes/no, if yes: description |
| 26 | EU | 20 Schemes to promote physical activity at the work place | Existence of a national or sub-national (where relevant#) incentive scheme for companies to promote physical activity at the work place (e.g. gyms, showers, walking stairs etc.): yes/no |
| 27 | EU | 21 Schemes for community interventions to promote PA in elderly people | Existence of a scheme for community interventions to promote PA in elderly people Yes/no, if yes: description |
| 28 | EU | 22 National HEPA policies that include a plan for evaluation | x out of y national HEPA policies (sport, health, transport, environment, by sector) include a clear intention or plan and plan for evaluation |
| 29 | EU | 23 Existence of a national awareness raising campaign on physical activity | Yes/no, if yes: description |
| 30 | GoPA | Tasks and subtasks | |
| 31 | GoPA | Budgets | |
| 32 | GoPA | Timelines-timeframe | |
| 33 | GoPA | Collaborators | |
| 34 | HEPA-PAT | 1 Background information and country context | 1a. Please provide a brief overview of the government structure in your country (about 200-400 words). For example, briefly outline whether your country has a centralized or federal system and on which government level the main responsibility lies for issues such as health, sport, |

| No. | Tool | Indicator / Category | Description |
|---|---|---|---|
| | | | education, transport, environment and urban planning policy. For examples relating to this and the other PAT questions, refer to the WHO website (www.euro.who.int/hepapat). |
| 35 | HEPA-PAT | 1 Background information and country context | 1b. Please briefly describe the governance at sub-national level (about 200-400 words) (e.g. at regional/provincial/cantonal/municipality level). |
| 36 | HEPA-PAT | 1 Background information and country context | 1c. Please provide a list of the main government ministries (e.g. health, sport, education, transport, environment and urban planning) in your national government that have a role in the promotion of HEPA (see Glossary for definition).<br>Please also include a brief description of the role(s) of these key HEPA-related government departments.<br>Please note: This question and Question 1d refer to the national level; please include information on the subnational level only where relevant, e.g. for countries with a strongly decentralized, federal system. |
| 37 | HEPA-PAT | 1 Background information and country context | 1d. Please list any other important national organizations, outside of government, which are actively engaged in HEPA promotion. This could include national sporting organizations, NGOs, charities, advocacy groups, the academic or scientific community, among others.<br>Please provide a brief description of the role of these organizations (about 50–100 words). |
| 38 | HEPA-PAT | 2 Leadership and partnerships | 2. Please state any agency(ies) providing leadership for HEPA promotion at the national level in your country. |
| 39 | HEPA-PAT | 2 Leadership and partnerships | 3. Please state any agency(ies) providing leadership for HEPA promotion at the subnational level (e.g. at regional/ provincial/cantonal/municipal level) in your country. |
| 40 | HEPA-PAT | 2 Leadership and partnerships | 4. Are any mechanisms or agencies in place in your country to ensure cross-sectoral collaboration on the delivery of HEPA policy, at the national level?<br>If yes, briefly describe. Please provide information on who is involved, who is leading these efforts, and how these collaborations function in practice. Please also mention (to the extent possible) any positive or more difficult experiences. This may also include examples of collaboration with the private and voluntary sectors. |
| 41 | HEPA-PAT | 2 Leadership and partnerships | 5. Are any mechanisms or bodies in place in your country to ensure cross-sectoral collaboration on the delivery of HEPA policy at the subnational level?<br>If yes, briefly describe. Please provide information on who is involved, who is leading these efforts, how these collaborations function in practice. Please also mention (to the extent possible) any positive or more difficult experiences. This may also include examples of collaboration with the private and voluntary sectors. |

| No. | Tool | Indicator / Category | Description |
|---|---|---|---|
| 42 | HEPA-PAT | 3 Policy documents | 6. Please describe any key past policy documents and past events that have led to the current context of HEPA promotion in your country. This might include legislation or recent policy documents that are now technically out of date (e.g. a previous national HEPA policy that may or may not have been extended), previous landmark legislation, or other documents such as scientific reports. Key events might include political changes, position statements or scientific events that have shaped the HEPA agenda.<br>Please list the documents/events, provide a web link (where available), and indicate if an English version or summary is available in each case. |
| 43 | HEPA-PAT | 3 Policy documents | 7. Please provide details (title, timeframe, issuing body) of the current key policy documents, legislation, strategies or action plans in your country, which outline government (and, where applicable, NGO) intention to increase national levels of physical activity (see Glossary for definitions of these terms).<br>Please list the documents according to sector and, where available, provide a web link, indicating whether an English version or summary is available. Please provide a brief description of the general content of each policy (about 100–250 words).<br>Please mark in the right-hand column which are the most important documents for the HEPA agenda in your country and briefly explain why these documents are deemed important.<br>Please add/remove rows as needed.<br>- Health<br>- Sport and recreation<br>- Education<br>- Transport<br>- Environment<br>- Urban design and planning<br>- Other sector |
| 44 | HEPA-PAT | 3 Policy documents | 8. During the development of the most important policies/action plans listed in Question 7, was a consultative process used, involving relevant stakeholders?<br>If yes, please briefly outline the steps of this consultation processes and which organizations were involved. Please also mention any challenges in recent years in engaging government ministries or other agencies through such processes. |
| 45 | HEPA-PAT | 3 Policy documents | 9. In your appraisal of the policy documents listed in Question 7, is there evidence of cross-referencing and alignment within and between policies, with genuine connections between |

| No. | Tool | Indicator / Category | Description |
|---|---|---|---|
| | | | different policy areas, or do the policies present separate, sector-specific strategies without evidence of links and consistency across sectors and documents with relevant policy?<br>For example: in the health sector, does a national obesity prevention strategy refer to an existing physical activity promotion plan, thus demonstrating an integrated overarching national approach to addressing physical activity? Does a transport policy recognize links with other policies that promote walking and cycling in the health sector (or sport field)? Does a sport promotion policy cross-reference HEPA promotion activities contained in a health promotion policy?<br>If yes, please briefly explain and give examples of such cross-referencing. Please state which of the policy documents presented in Question 7 you are referring to. |
| 46 | HEPA-PAT | 3 Policy documents | 10. In your country, are any mechanisms in place to ensure that the key policy documents listed in Question 7 are based on the best-available scientific evidence on HEPA?<br>For example, are specific mechanisms or agencies dedicated to reviewing evidence and ensuring that the latest evidence is used to inform national policy development? Do any formal committees or institutions exist that are responsible for reviewing evidence and providing guidance to national policy-making bodies, or any formal links between government and academic institutions for this purpose?<br>If yes, please briefly describe these. |
| 47 | HEPA-PAT | 3 Policy documents | 11. Please indicate how useful the following international documents have been in the development of physical activity- related policy in your country, e.g. by serving as a basis, input or inspiration (whether having been specifically quoted or not in a policy document). Please rate the documents below on the scale from 1 (= “not at all useful”) to 5 (= “very useful”). Please add any other international documents which have been important in the development of physical activity-related policy in your country, as necessary.<br><br>Global strategy on diet, physical activity and health (2)<br>Global recommendations on physical activity for health (3)<br>2008–2013 action plan for the global strategy for the prevention and control of noncommunicable diseases (4)<br>Global status report on noncommunicable diseases 2010 (5)<br>Global action plan for the prevention and control of noncommunicable diseases 2013–2020 (6)<br>Steps to health. A European framework to promote physical activity for health (7) |

| No. | Tool | Indicator / Category | Description |
|---|---|---|---|
| | | | Action plan for implementation of the European strategy for the prevention and control of noncommunicable diseases 2012−2016 (8)<br>The Toronto Charter for physical activity: a global call for action (9)<br>Noncommunicable disease prevention: investments that work for physical activity (10)<br>Lancet series on Physical Activity (11)<br>Other document (please specify): |
| 48 | HEPA-PAT | 3 Policy documents | 12. Do any national documents or guidelines exist that support implementation of HEPA activities at the subnational level? For example, does national policy determine what is delivered at the subnational level and, if so, is this national guidance strongly adhered to? Such guidance could include programmes, structures or funding. Or is subnational policy and activity developed and implemented largely independently from the national government?<br>Please note: please be brief here (about 300–500 words) and include cross-references to other questions (e.g. Question 7) where relevant, to avoid repetition. |
| 49 | HEPA-PAT | 4 Policy scope, content and implementation | 13. Considering all the key physical activity policy documents listed in Question 7, please indicate which settings are included for the delivery of specific HEPA actions.<br>Please only tick those settings in which dedicated programmes or interventions are foreseen or already under way.<br><br>Preschools/kindergarten<br>Primary schools<br>Secondary/high schools<br>Colleges/universities<br>Primary health care<br>Clinical health care (e.g. hospitals)<br>Workplace<br>Older adult/senior services<br>Sport and recreation<br>Transport<br>Tourism<br>Environment<br>Urban design and planning |

| No. | Tool | Indicator / Category | Description |
| --- | --- | --- | --- |
| | | | Community<br>Other (please specify) |
| 50 | HEPA-PAT | 4 Policy scope, content and implementation | 14. Considering all the key physical activity policy documents listed in Question 7, please indicate which population groups are targeted by specific HEPA actions.<br>Please only tick those groups for which dedicated programmes or interventions are foreseen or already under way.<br><br>Early years<br>Children/young people<br>Older adults<br>Workforce/employees<br>Women<br>People with disabilities<br>Clinical populations/chronic disease patients<br>Sedentary/the least active<br>People from low socio-economic status<br>Families<br>Indigenous people<br>Migrant populations<br>General population<br>Other (please specify): |
| 51 | HEPA-PAT | 4 Policy scope, content and implementation | 15. Does your country have a current national communication strategy (using mass media) aimed at raising awareness and promoting physical activity?<br>If yes, please provide details of the communication activities (e.g. posters, website, television or radio advertising, etc.) and whether these activities have a common branding or slogan (e.g. “Agita Sao Paulo” or “Find 30”).<br>If no, has your country conducted any national communication activities in the past? |
| 52 | HEPA-PAT | 4 Policy scope, content and implementation | 16. To illustrate the types of policy actions in your country, please provide one or two examples (if available) of large-scale (preferably national) programmes or interventions in each of the settings listed.<br>Please provide a brief description of each programme or intervention (about 100 words, including, for example: name, lead organization, approach, participants, results.) and a source |

| No. | Tool | Indicator / Category | Description |
|---|---|---|---|
| | | | where further information can be obtained.<br>Suggestion: You could also consider developing these examples into more detailed case studies to complement your national PAT assessment.<br><br>Health<br>Sport/recreation<br>Education<br>Transport<br>Environment<br>Urban design/planning<br>Other |
| 53 | HEPA-PAT | 5 Recommendations, goals and targets | 17a. Does your country have any national recommendations on physical activity and health?<br>National recommendations refer to a consensus statement on how much activity is required for health benefits.<br>If recommendations exist for any of the target groups listed, please provide details for the population subgroups (where applicable), including issuing body, year of publication, title of the document, and provide a web link if available (please also specify whether the document is available in English).<br>If no recommendations exist, please mark the "no" column for the respective target group. If your country has officially adopted or endorsed international recommendations (e.g. of WHO or the United States Department of Health), this should be mentioned as part of the description of the respective recommendations.<br><br>Early years (pre-school age)<br>Children and young people (school-age)<br>Adults<br>Older adults/seniors<br>People with disabilities<br>Other (please specify): |
| 54 | HEPA-PAT | 5 Recommendations, goals and targets | 17b. Does your country have any national recommendations on reducing sedentary behaviour?<br>If recommendations exist for any of the target groups listed, please provide details for each of the population subgroups (where applicable), including the issuing body, year of publication, title |

| No. | Tool | Indicator / Category | Description |
| --- | --- | --- | --- |
| | | | of the document, and provide a web link if available (please also specify whether the document is available in English).<br>If no recommendations exist, please mark the “no” column for the respective target group.<br><br>Early years (pre-school age)<br>Children and young people (school-age)<br>Adults<br>Older adults/seniors<br>People with disabilities<br>Other (please specify): |
| 55 | HEPA-PAT | 5 Recommendations, goals and targets | 18. Does your country have any national goals (or national targets) for population prevalence of physical activity? If yes, please provide details of each target and the time frame. Please specify in which policy document(s) listed in Question 7 these goals are stated.<br>Please start with the most specific and measurable targets, followed by a listing or summary statement of any more general targets and goals for physical activity-related behaviours. |
| 56 | HEPA-PAT | 5 Recommendations, goals and targets | 19. Aside from any national goals and targets for population prevalence of physical activity or sedentary behaviour (already provided in previous questions), does your country have any other goals and targets that directly or indirectly relate to physical activity promotion? |
| 57 | HEPA-PAT | 6 Surveillance | 20. Does your country have a health surveillance or monitoring system that includes measures of physical activity or sedentary behaviour?<br>If yes, please provide details according to age group (you may copy and paste as many response sections as needed). Please describe long-term general population surveys in: Question 20a (children and young people); Question 20b (adults) and Question 20c (older adults/seniors). Please add more boxes if needed.<br><br>20a. Children and young people (name of survey, methods used, part of repeated surveillance system, single survey(s))<br>20b. Adults (name of survey, methods used, part of repeated surveillance system, single survey(s))<br>20c. Older adults (name of survey, methods used, part of repeated surveillance system, single survey(s)) |

| No. | Tool | Indicator / Category | Description |
| --- | --- | --- | --- |
| 58 | HEPA-PAT | 6 Surveillance | 21a. Have data on the prevalence of physical activity or sedentary behaviour or other related factors influenced policy development in your country?<br>For example, have surveillance data been used to define national goals and targets, or to assess progress towards achieving national goals and targets? If yes, please explain briefly and give examples.<br>If no, please briefly explain why. For example, is the frequency of data collection not in line with the timeline of formulated policy goals, or do the questions asked in the survey not provide information on the effectiveness of national policy implementation? |
| 59 | HEPA-PAT | 6 Surveillance | 21b. In your opinion, have surveillance data helped to progress the national promotion of physical activity in your country in any other ways?<br>For example, has a decline of physical activity levels helped to increase political attention, or created media attention?<br>If yes, please explain briefly, giving examples. |
| 60 | HEPA-PAT | 7 Evaluation | 22a. Has your country undertaken evaluation of any of the national policies or action plans listed in Question 7?<br>If yes, please state the title of the report, publisher and year published. Where available, please also provide a web link and indicate whether an English version/summary is available. Please provide brief details of the evaluation undertaken, what has been evaluated, the data collection methods, a summary of the results and how these were used (or not) to define new policy.<br><br>Title<br>Publisher and date<br>Web link<br>Brief description of the approaches, results and their use |
| 61 | HEPA-PAT | 7 Evaluation | 22b. Has any evaluation of physical activity projects or interventions taken place at the subnational level (coordinated with or independent from the national level)?<br>If yes, please give a brief general overview of relevant processes. It is not expected to cover the whole range of activities but rather to give an indication and overview of the general approach taken at the subnational level. |
| 62 | HEPA-PAT | 7 Evaluation | 23. Has any economic evaluation of interventions or physical inactivity (i.e. not reaching the minimum recommended level of physical activity) at national level been undertaken in your country? |

| No. | Tool | Indicator / Category | Description |
|---|---|---|---|
| | | | If yes, please state the title of the report, publisher and year published. Where available, please also provide a web link and indicate whether an English version/summary is available. Please provide a brief description of the results of the assessment (about 50–100 words).<br><br>Title<br>Publisher and date<br>Web link<br>Brief description of the approaches, results and their use |
| 63 | HEPA-PAT | 8 Funding and commitments | 24a. Within each of the sectors listed, is funding specifically allocated or "ring-fenced" for the delivery of physical activity- related policy or action plans at the national level?<br>Please tick yes/no, and provide the amount (and currency), if known. Please also indicate whether this funding is recurrent; that is, provided on a regular basis (e.g. annually).<br><br>Health (yes, amount, no, don't know - recurrent: yes, no, don't know)<br>Sport/recreation<br>Education<br>Transport<br>Environment<br>Urban design/planning<br>Other (please specify) |
| 64 | HEPA-PAT | 8 Funding and commitments | 24b. Within each of the sectors listed, is funding specifically allocated or "ring-fenced" for the delivery of physical activity- related policy or action plans at the subnational level?<br>Please tick yes/no, and provide the amount (and currency), if known. Please also indicate whether this funding is recurrent; that is, provided on a regular basis (e.g. annually).<br><br>Health (yes, amount, no, don't know - recurrent: yes, no, don't know)<br>Sport/recreation<br>Education<br>Transport<br>Environment |

| No. | Tool | Indicator / Category | Description |
|---|---|---|---|
| | | | Urban design/planning<br>Other (please specify) |
| 65 | HEPA-PAT | 8 Funding and commitments | 25. In your opinion, does evidence exist of political commitment to the national agenda to promote physical activity? This might include, for example: recognition of physical activity as an important policy topic; increased funding; inclusion of physical activity in official speeches; political discussions about physical activity promotion in parliament; visible engagement by politicians in HEPA-related events, or their personal participation in HEPA.<br>If yes, please describe, giving examples, and also comment on whether you think there is greater or less political commitment to physical activity promotion in your country than in the recent past. |
| 66 | HEPA-PAT | 9 Capacity-building through a national network | 26. Does any professional network or system exist in your country that links and/or supports professionals interested or currently working in physical activity or related areas?<br>If yes, please describe, providing a web link and contact person, where available. |
| 67 | HEPA-PAT | 10 Experience of policy implementation, progress and remaining challenges | 27a. What do you think are the areas of greatest progress in national HEPA promotion in your country in recent years? |
| 68 | HEPA-PAT | 10 Experience of policy implementation, progress and remaining challenges | 27b. What do you think have been the biggest challenges faced by your country in the commencement or continuation of national-level approaches to HEPA promotion in recent years? |
| 69 | HEPA-PAT | 10 Experience of policy implementation, progress and remaining challenges | 28. Based on your experience, please identify up to three suggestions you would offer to another country that is setting up a national HEPA policy. |
| 70 | HEPA-PAT | 10 Experience of policy implementation, progress and remaining challenges | 29. Please use this space to provide any further details or comments you were not able to provide in other sections of the tool. |
| 71 | INTEGRATE | A. National level PA policy | Please provide detailed information about the current national physical activity policy that is being<br>implemented for [COUNTRY] [a1_c1]<br><br>For the purposes of this survey, physical activity policy is understood as those formally written policies, rules and guidelines, written non-formal direct statements, formal procedures and informal policies (or lack thereof) that may or indirectly affect physical activity through |

| No. | Tool | Indicator / Category | Description |
|---|---|---|---|
| | | | community or population level.<br>Consider that "the promotion of physical activity is not directly named", for example, that the document mentions active transport, sustainable transport, recreation, sports, cycling, etc.<br><br>[title_c1] National Policy Title:<br>[pubyr_c1] Publication year:<br>[timefr_c1] Time frame covered:<br>[issuing_c1] Issuing body:<br>[weblink_c1] Web link: |
| 72 | INTEGRATE | A. National level PA policy | 2. Is the policy standalone for physical activity promotion? [excl_c1]<br>Yes ☐ [1] [Go to 3]<br>No ☐ [0] [Go to 2.b]<br>I don't know ☐ [99] |
| 73 | INTEGRATE | A. National level PA policy | 2.b. Does the policy include physical activity promotion as part of another policy or plan, whose focus is not exclusively on physical activity? [otherplan_c1]<br>Yes ☐ [1] [Go to 2.b.1]<br>No ☐ [0] [Go to 3]<br>I don't know ☐ [99] |
| 74 | INTEGRATE | A. National level PA policy | 2.b.1 If yes, in what type of policy or plan is it embedded? [othertype_c1]<br>☐ NCDs prevention plan [1]<br>☐ Obesity prevention and/or management or control plan [2]<br>☐ A plan not related to public health (e.g., sports, environment, etc.) [3]<br>· Specify the other type of policy or plan: __________ [type_sp_c1] |
| 75 | INTEGRATE | A. National level PA policy | 3. In this policy, what role does the promotion of physical activity play? [afrole_c1]<br>☐ Central (e.g., it is an exclusive policy for physical activity) [1]<br>☐ Important but not central (e.g., physical activity is included in another policy or plan) [2]<br>☐ Secondary or perhaps physical activity is not mentioned; however, the document has an influence on physical activity [3] |
| 76 | INTEGRATE | B. Policy Operational structure (About regional policymakers) | **1. Agenda Setting Stage [reg_nat_agenda_c1]**<br>Refers to the process through which a problem is identified and structured as of public interest and is intended to be addressed through public policy. |

| No. | Tool | Indicator / Category | Description |
|---|---|---|---|
| | | | Thinking about the agenda setting stage for the development of national physical activity policies....<br><br>1.a. To the best of your knowledge, are representatives of the regional health sectors usually invited to participate in the agenda setting stage for the development of national physical activity policies? [reg_nat_agenda2_c1]<br>Yes ☐ [1] [Go to 1.b.]<br>No ☐ [0]<br>I don't know ☐ [99]<br><br>1.b. How would you rate the level of involvement of regional health sectors in the agenda setting stage for<br>developing national physical activity policies? [reg_nat_agenda2a_c1]<br>No involvement ☐[0] ☐[1] ☐[2] ☐[3] ☐[4] ☐[5] Extensive involvement; I don't know ☐[99]<br><br>1.c. (OPTIONAL) – Could you share more information on how regional health sectors influence or participate in the agenda setting stage for the development of national physical activity policies? [Open Ended Question] [reg_nat_agenda3_c1]<br>________________________________________<br>________________________________________ |
| 77 | INTEGRATE | B. Policy Operational structure (About regional policymakers) | 2. Policy Formulation Stage [reg_nat_form_c1]<br>Refers to the process in which public administration examines the different policy options that can be considered as possible solutions for the problem.<br>Thinking about the policy formulation stage for the development of national physical activity policies....<br><br>2.a. To the best of your knowledge, are representatives of the regional health sectors usually invited to participate in the policy formulation stage for the development of national physical activity policies? [reg_nat_form2_c1]<br>Yes ☐ [1] [Go to 2.b.]<br>No ☐ [0] |

| No. | Tool | Indicator / Category | Description |
|---|---|---|---|
| | | | I don't know ☐ [99]<br><br>2.b. How would you rate the level of involvement of regional health sectors in the policy formulation stage for developing national physical activity policies? [reg_nat_form2a_c1]<br>No involvement ☐[0] ☐[1] ☐[2] ☐[3] ☐[4] ☐[5] Extensive involvement; I don't know ☐[99]<br><br>2.c. (OPTIONAL) – Could you share more information on how regional health sectors influence or participate in the policy formulation stage for the development of national physical activity policies? [Open Ended Question] [reg_nat_form3_c1]<br>____________________________________________________________<br>____________________________________________________________ |
| 78 | INTEGRATE | B. Policy Operational structure (About regional policymakers) | 3. Adoption (decision making) Stage [reg_nat_adop_c1]<br>The stage during which decisions occur at a governmental level, resulting in selecting solutions for the policy<br>Thinking about the adoption (decision making) stage for the development of national physical activity policies...<br><br>3.a. To the best of your knowledge, are representatives of the regional health sectors usually invited to participate in the adoption (decision making) stage for the development of national physical activity policies? [reg_nat_adop2_c1]<br>Yes ☐ [1] [Go to 3.b.]<br>No ☐ [0]<br>I don't know ☐ [99]<br><br>3.b. How would you rate the level of involvement of regional health sectors in the adoption (decision<br>making) stage for developing national physical activity policies? [reg_nat_adop2a_c1]<br>No involvement ☐[0] ☐[1] ☐[2] ☐[3] ☐[4] ☐[5] Extensive involvement; I don't know ☐[99]<br><br>3.c. (OPTIONAL) – Could you share more information on how regional health sectors influence or participate in the adoption (decision making) stage for the development of national physical |

| No. | Tool | Indicator / Category | Description |
|---|---|---|---|
| | | | activity policies? [Open Ended Question] [reg_nat_adop3_c1]<br>______________________________________________________________________<br>______________________________________________________________________ |
| 79 | INTEGRATE | B. Policy Operational structure (About regional policymakers) | 4. Implementation stage [reg_nat_impl_c1]<br>Process in which the parameters for implementing the policy are established. At this same stage, objectives are set.<br>Thinking about the implementation stage for the development of national physical activity policies....<br><br>4.a. To the best of your knowledge, are representatives of the regional health sectors usually participating in the implementation stage of national physical activity policies? [reg_nat_impl2_c1]<br>Yes ☐ [1] [Go to 4.b.] No ☐ [0] I don't know ☐ [99]<br><br>4.b. How would you rate the level of involvement of regional health sectors in the implementation stage for developing national physical activity policies? [reg_nat_impl2a_c1]<br>No involvement ☐[0] ☐[1] ☐[2] ☐[3] ☐[4] ☐[5] Extensive involvement; I don't know ☐[99]<br><br>4.c. (OPTIONAL) – Could you share more information on how regional health sectors influence or participate in the implementation stage for the development of national physical activity policies? [Open Ended Question] [reg_nat_impl3_c1]<br>______________________________________________________________________<br>______________________________________________________________________<br>4.d. Is there coordination between the national level and the regional level health sectors during the implementation process of national physical activity policies? [reg_nat_impl4_c1]<br>Yes ☐ [1] No ☐ [0] I don't know ☐ [99] |
| 80 | INTEGRATE | B. Policy Operational structure (About regional policymakers) | 5. Evaluation stage [reg_nat_eval_c1]<br>Process in which a policy is evaluated to assess its effectiveness concerning the policy objectives.<br>Thinking about the evaluation stage for the development of national physical activity policies....<br><br>5.a. To the best of your knowledge, are representatives of the regional health sectors usually invited to participate in the evaluation stage of national physical activity policies? |

| No. | Tool | Indicator / Category | Description |
|---|---|---|---|
| | | | [reg_nat_eval2_c1]<br>Yes ☐ [1] [Go to 5.b.] No ☐ [0] I don't know ☐ [99]<br><br>5.b. How would you rate the level of involvement of regional health sectors in the evaluation stage for<br>developing national physical activity policies? [reg_nat_eval2a_c1]<br>No involvement ☐[0] ☐[1] ☐[2] ☐[3] ☐[4] ☐[5] Extensive involvement; I don't know ☐[99]<br><br>5.c. (OPTIONAL) – Could you share more information on how regional health sectors influence or participate in the evaluation stage for the development of national physical activity policies? [Open Ended Question] [reg_nat_eval3_c1]<br>__________________________________________________<br>__________________________________________________<br><br>5.d. Is there a formal (e.g., legally binding or a policy statement) process to evaluate the work (priorities, responsibilities, implementation) related to national physical activity policies between the national level and the regional level health sectors? [reg_nat_eval4_c1]<br>Yes ☐ [1] No ☐ [0] I don't know ☐ [99] |
| 81 | INTEGRATE | B. Policy Operational structure<br>(About subnational policymakers) | 1. Agenda Setting Stage [nat_agenda_c1]<br>Refers to the process through which a problem is identified and structured as of public interest and is intended to be addressed through public policy.<br>Thinking about the agenda setting stage for the development of national physical activity policies....<br><br>1.a. To the best of your knowledge, are representatives of the subnational (city-level) health sectors usually invited to participate in the agenda setting stage for the development of national physical activity policies? [nat_agenda2_c1]<br>Yes ☐ [1] [Go to 1.b.]<br>No ☐ [0]<br>I don't know ☐ [99] |

| No. | Tool | Indicator / Category | Description |
|---|---|---|---|
| | | | 1.b. How would you rate the level of involvement of subnational (city-level) health sectors in the agenda setting stage for developing national physical activity policies? [nat_agenda2a_c1]<br>No involvement ☐[0] ☐[1] ☐[2] ☐[3] ☐[4] ☐[5] Extensive involvement; I don't know ☐[99]<br><br>1.c. (OPTIONAL) – Could you share more information on how subnational (city-level) health sectors influence or participate in the agenda setting stage for the development of national physical activity policies? [Open Ended Question] [nat_agenda3_c1]<br>_______________________________________________________________________<br>_______________________________________________________________________ |
| 82 | INTEGRATE | B. Policy Operational structure (About subnational policymakers) | 2. Policy Formulation Stage [nat_form_c1]<br>Refers to the process in which public administration examines the different policy options that can be considered as possible solutions for the problem.<br>Thinking about the policy formulation stage for the development of national physical activity policies....<br><br>2.a. To the best of your knowledge, are representatives of the subnational (city-level) health sectors usually invited to participate in the policy formulation stage for the development of national physical activity policies? [nat_form2_c1]<br>Yes ☐ [1] [Go to 2.b.]<br>No ☐ [0]<br>I don't know ☐ [99]<br><br>2.b. How would you rate the level of involvement of subnational (city-level) health sectors in the policy formulation stage for developing national physical activity policies? [nat_form2a_c1]<br>No involvement ☐[0] ☐[1] ☐[2] ☐[3] ☐[4] ☐[5] Extensive involvement; I don't know ☐[99]<br><br>2.c. (OPTIONAL) – Could you share more information on how subnational (city-level) health sectors influence or participate in the policy formulation stage for the development of national physical activity policies? [Open Ended Question] [nat_form3_c1]<br>_______________________________________________________________________<br>_______________________________________________________________________ |

| No. | Tool | Indicator / Category | Description |
|---|---|---|---|
| 83 | INTEGRATE | B. Policy Operational structure<br>(About subnational policymakers) | 3. Adoption (decision making) Stage [nat_adop_c1]<br>The stage during which decisions occur at a governmental level, resulting in selecting solutions for the policy<br>Thinking about the adoption (decision making) stage for the development of national physical activity policies...<br><br>3.a. To the best of your knowledge, are representatives of the subnational (city-level) health sectors usually invited to participate in the adoption (decision making) stage for the development of national physical activity policies? [nat_adop2_c1]<br>Yes ☐ [1] [Go to 3.b.]<br>No ☐ [0]<br>I don't know ☐ [99]<br><br>3.b. How would you rate the level of involvement of subnational (city-level) health sectors in the adoption (decision making) stage for developing national physical activity policies? [nat_adop2a_c1]<br>No involvement ☐[0] ☐[1] ☐[2] ☐[3] ☐[4] ☐[5] Extensive involvement; I don't know ☐[99]<br><br>3.c. (OPTIONAL) – Could you share more information on how subnational (city-level) health sectors influence or participate in the adoption (decision making) stage for the development of national physical activity policies? [Open Ended Question] [nat_adop3_c1]<br>______________________________________________________________________________<br>______________________________________________________________________________ |
| 84 | INTEGRATE | B. Policy Operational structure<br>(About subnational policymakers) | 4. Implementation stage [nat_impl_c1]<br>Process in which the parameters for implementing the policy are established. At this same stage, objectives are set.<br>Thinking about the implementation stage for the development of national physical activity policies....<br><br>4.a. To the best of your knowledge, are representatives of the subnational (city-level) health sectors usually participating in the implementation stage of national physical activity policies? [nat_impl2_c1] |

| No. | Tool | Indicator / Category | Description |
|---|---|---|---|
| | | | Yes ☐ [1] [Go to 4.b.] No ☐ [0] I don't know ☐ [99]<br><br>4.b. How would you rate the level of involvement of subnational (city-level) health sectors in the implementation stage for developing national physical activity policies? [nat_impl2a_c1]<br>No involvement ☐[0] ☐[1] ☐[2] ☐[3] ☐[4] ☐[5] Extensive involvement; I don't know ☐[99]<br><br>4.c. (OPTIONAL) – Could you share more information on how subnational (city-level) health sectors influence or participate in the implementation stage for the development of national physical activity policies? [Open Ended Question] [nat_impl3_c1]<br>________________________________________________<br>________________________________________________<br>4.d. Is there coordination between the national level and the subnational (city-level) level health sectors during the implementation process of national physical activity policies? [nat_impl4_c1]<br>Yes ☐ [1] No ☐ [0] I don't know ☐ [99] |
| 85 | INTEGRATE | B. Policy Operational structure (About subnational policymakers) | 5. Evaluation stage [nat_eval_c1]<br>Process in which a policy is evaluated to assess its effectiveness concerning the policy objectives. Thinking about the evaluation stage for the development of national physical activity policies....<br><br>5.a. To the best of your knowledge, are representatives of the subnational (city-level) health sectors usually invited to participate in the evaluation stage of national physical activity policies? [nat_eval2_c1]<br>Yes ☐ [1] [Go to 5.b.] No ☐ [0] I don't know ☐ [99]<br><br>5.b. How would you rate the level of involvement of subnational (city-level) health sectors in the evaluation stage for developing national physical activity policies? [nat_eval2a_c1]<br>No involvement ☐[0] ☐[1] ☐[2] ☐[3] ☐[4] ☐[5] Extensive involvement; I don't know ☐[99]<br><br>5.c. (OPTIONAL) – Could you share more information on how subnational (city-level) health sectors influence or participate in the evaluation stage for the development of national physical activity policies? [Open Ended Question] [nat_eval3_c1]<br>________________________________________________ |

| No. | Tool | Indicator / Category | Description |
|---|---|---|---|
| | | | ________________________________________________________________________________<br><br>5.d. Is there a formal (e.g., legally binding or a policy statement) process to evaluate the work (priorities, responsibilities, implementation) related to national physical activity policies between the national level and the subnational (city-level) level health sectors? [nat_eval4_c1]<br>Yes ☐ [1] No ☐ [0] I don't know ☐ [99] |
| 86 | INTEGRATE | B. Policy operational structure (Regional level policies) | 1. Agenda Setting Stage [reg_agenda_c1]<br>Refers to the process through which a problem is identified and structured as of public interest and is intended to be addressed through public policy.<br>Thinking about the agenda setting stage for the development of regional physical activity policies....<br><br>1.a. To the best of your knowledge, are representatives of the national health sector usually invited to participate in the agenda setting stage for the development of regional physical activity policies? [reg_agenda2_c1]<br>Yes ☐ [1] [Go to 1.b.]<br>No ☐ [0]<br>I don't know ☐ [99]<br><br>1.b. How would you rate the level of involvement of the national health sector in the agenda setting stage for developing regional physical activity policies? [reg_agenda2a_c1]<br>No involvement ☐[0] ☐[1] ☐[2] ☐[3] ☐[4] ☐[5] Extensive involvement; I don't know ☐[99]<br><br>1.c. (OPTIONAL) – Could you share more information on how the national health sector influences or participates in the agenda setting stage for the development of regional physical activity policies? [Open Ended Question] [reg_agenda3_c1]<br>________________________________________________________________________________<br>________________________________________________________________________________ |
| 87 | INTEGRATE | B. Policy operational structure (Regional level policies) | 2. Policy Formulation Stage [reg_form_c1]<br>Refers to the process in which public administration examines the different policy options that can be considered as possible solutions for the problem. |

| No. | Tool | Indicator / Category | Description |
|---|---|---|---|
| | | | Thinking about the policy formulation stage for the development of regional physical activity policies....<br><br>2.a. To the best of your knowledge, are representatives of the national health sector usually invited to participate in the policy formulation stage of regional physical activity policies? [reg_form2_c1]<br>Yes ☐ [1] [Go to 2.b.]<br>No ☐ [0]<br>I don’t know ☐ [99]<br><br>2.b. How would you rate the level of involvement of the national health sector in the policy formulation stage of regional physical activity policies? [reg_form2a_c1]<br>No involvement ☐[0] ☐[1] ☐[2] ☐[3] ☐[4] ☐[5] Extensive involvement; I don't know ☐[99]<br><br>2.c. (OPTIONAL) – Could you share more information on how the national health sector influences or participates in the policy formulation stage for the development of regional physical activity policies? [Open Ended Question] [reg_form3_c1]<br>________________________________________<br>________________________________________ |
| 88 | INTEGRATE | B. Policy operational structure (Regional level policies) | 3. Adoption (decision making) Stage [reg_adop_c1]<br>The stage during which decisions occur at a governmental level, resulting in selecting solutions for the policy<br>Thinking about the adoption (decision making) stage for the development of regional physical activity policies...<br><br>3.a. To the best of your knowledge, are representatives of the national health sector usually invited to participate in the adoption (decision making) stage of regional physical activity policies? [reg_adop2_c1]<br>Yes ☐ [1] [Go to 3.b.]<br>No ☐ [0]<br>I don’t know ☐ [99] |

| No. | Tool | Indicator / Category | Description |
|---|---|---|---|
| | | | 3.b. How would you rate the level of involvement of the national health sector in the adoption (decision<br>making) stage for developing regional physical activity policies? [reg_adop2a_c1]<br>No involvement ☐[0] ☐[1] ☐[2] ☐[3] ☐[4] ☐[5] Extensive involvement; I don't know ☐[99]<br><br>3.c. (OPTIONAL) – Could you share more information on how the national health sector influences or participates in the adoption (decision making) stage for the development of regional physical activity policies? [Open Ended Question] [reg_adop3_c1]<br>____________________________________________________________<br>____________________________________________________________ |
| 89 | INTEGRATE | B. Policy operational structure (Regional level policies) | 4. Implementation stage [reg_impl_c1]<br>Process in which the parameters for implementing the policy are established. At this same stage, objectives are set.<br>Thinking about the implementation stage for the development of regional physical activity policies....<br><br>4.a. To the best of your knowledge, are representatives of the national health sector usually participating in the implementation stage of regional physical activity policies? [reg_impl2_c1]<br>Yes ☐ [1] [Go to 4.b.] No ☐ [0] I don't know ☐ [99]<br><br>4.b. How would you rate the level of involvement of the national health sector in the implementation stage for developing regional physical activity policies? [reg_impl2a_c1]<br>No involvement ☐[0] ☐[1] ☐[2] ☐[3] ☐[4] ☐[5] Extensive involvement; I don't know ☐[99]<br><br>4.c. (OPTIONAL) – Could you share more information on how the national health sector influences or participates in the implementation stage for the development of regional physical activity policies? [Open Ended Question] [reg_impl3_c1]<br>____________________________________________________________<br>____________________________________________________________<br>4.d. Is there coordination between the national level and the regional level health sectors during |

| No. | Tool | Indicator / Category | Description |
|---|---|---|---|
| | | | the implementation process of regional physical activity policies? [reg_impl4_c1]<br>Yes ☐ [1] No ☐ [0] I don't know ☐ [99] |
| 90 | INTEGRATE | B. Policy operational structure (Regional level policies) | 5. Evaluation stage [reg_nat_eval_c1]<br>Process in which a policy is evaluated to assess its effectiveness concerning the policy objectives. Thinking about the evaluation stage for the development of regional physical activity policies....<br><br>5.a. To the best of your knowledge, are representatives of the national health sector usually participating in the evaluation stage of regional physical activity policies? [reg_eval2_c1]<br>Yes ☐ [1] [Go to 5.b.] No ☐ [0] I don't know ☐ [99]<br><br>5.b. How would you rate the level of involvement of the national health sector in the evaluation stage of regional physical activity policies? [reg_eval2a_c1]<br>No involvement ☐[0] ☐[1] ☐[2] ☐[3] ☐[4] ☐[5] Extensive involvement; I don't know ☐[99]<br><br>5.c. (OPTIONAL) – Could you share more information on how the national health sector influences or participates in the evaluation stage for the development of regional physical activity policies? [Open Ended Question] [reg_eval3_c1]<br>____________________________________________________________<br>____________________________________________________________<br><br>5.d. Is there a formal (e.g., legally binding or a policy statement) process to evaluate the work (priorities, responsibilities, implementation) related to regional physical activity policies between the national level and the regional level health sectors? [reg_eval4_c1]<br>Yes ☐ [1] No ☐ [0] I don't know ☐ [99] |
| 91 | INTEGRATE | B. Policy operational structure (Regional level policies) | 6. To what extent do you consider your office (national health sector) to be involved in agenda setting, policy formulation, adoption (decision making), implementation and evaluation of regional physical activity policies for the following cities? [part_desc_c1] Please review the survey invitation email, which specifies for which cities you are responding to this question.<br>Region 1: No involvement ☐[0] ☐[1] ☐[2] ☐[3] ☐[4] ☐[5] Extensive involvement; I don't know ☐[99] |

| No. | Tool | Indicator / Category | Description |
|---|---|---|---|
| 92 | INTEGRATE | B. Policy operational structure (Subnational city-level policies) | 1. Agenda Setting Stage [agenda_c1]<br>Refers to the process through which a problem is identified and structured as of public interest and is intended to be addressed through public policy.<br>Thinking about the agenda setting stage for the development of subnational (city-level) physical activity policies....<br><br>1.a. To the best of your knowledge, are representatives of the national health sector usually participating in the agenda setting stage for the development of subnational (city-level) physical activity policies? [agenda2_c1]<br>Yes ☐ [1] [Go to 1.b.]<br>No ☐ [0]<br>I don't know ☐ [99]<br><br>1.b. How would you rate the level of involvement of the national health sector in the agenda setting stage for developing subnational (city-level) physical activity policies? [agenda2a_c1]<br>No involvement ☐[0] ☐[1] ☐[2] ☐[3] ☐[4] ☐[5] Extensive involvement; I don't know ☐[99]<br><br>1.c. (OPTIONAL) – Could you share more information on how the national health sector influences or participates in the agenda setting stage for the development of subnational (city-level) physical activity policies? [Open Ended Question] [agenda3_c1]<br>__________________________________________________________________________<br>__________________________________________________________________________ |
| 93 | INTEGRATE | B. Policy operational structure (Subnational city-level policies) | 2. Policy Formulation Stage [form_c1]<br>Refers to the process in which public administration examines the different policy options that can be considered as possible solutions for the problem.<br>Thinking about the policy formulation stage for the development of subnational (city-level) physical activity policies....<br><br>2.a. To the best of your knowledge, are representatives of the national health sector usually participating in the policy formulation stage of subnational (city-level) physical activity policies? [form2_c1] |

| No. | Tool | Indicator / Category | Description |
|---|---|---|---|
| | | | Yes ☐ [1] [Go to 2.b.]<br>No ☐ [0]<br>I don’t know ☐ [99]<br><br>2.b. How would you rate the level of involvement of the national health sector in the policy formulation stage for developing subnational (city-level) physical activity policies? [form2a_c1]<br>No involvement ☐[0] ☐[1] ☐[2] ☐[3] ☐[4] ☐[5] Extensive involvement; I don't know ☐[99]<br><br>2.c. (OPTIONAL) – Could you share more information on how the national health sector influences or participates in the policy formulation stage for the development of subnational (city-level) physical activity policies? [Open Ended Question] [form3_c1]<br>________________________________________________________________________________<br>________________________________________________________________________________ |
| 94 | INTEGRATE | B. Policy operational structure (Subnational city-level policies) | 3. Adoption (decision making) Stage [adop_c1]<br>The stage during which decisions occur at a governmental level, resulting in selecting solutions for the policy<br>Thinking about the adoption (decision making) stage for the development of subnational (city-level) physical activity policies...<br><br>3.a. To the best of your knowledge, are representatives of the national health sector usually participating in the adoption (decision making) stage of subnational (city-level) physical activity policies? [adop2_c1]<br>Yes ☐ [1] [Go to 3.b.]<br>No ☐ [0]<br>I don’t know ☐ [99]<br><br>3.b. How would you rate the level of involvement of the national health sector in the adoption (decision<br>making) stage for developing subnational (city-level) physical activity policies? [adop2a_c1]<br>No involvement ☐[0] ☐[1] ☐[2] ☐[3] ☐[4] ☐[5] Extensive involvement; I don't know ☐[99] |

| No. | Tool | Indicator / Category | Description |
|---|---|---|---|
| | | | 3.c. (OPTIONAL) – Could you share more information on how the national health sector influences or participates in the adoption (decision making) stage for the development of subnational (city-level) physical activity policies? [Open Ended Question] [adop3_c1]<br>________________________________________<br>________________________________________ |
| 95 | INTEGRATE | B. Policy operational structure (Subnational city-level policies) | 4. Implementation stage [impl_c1]<br>Process in which the parameters for implementing the policy are established. At this same stage, objectives are set.<br>Thinking about the implementation stage for the development of subnational (city-level) physical activity policies....<br><br>4.a. To the best of your knowledge, are representatives of the national health sector usually participating in the implementation stage of subnational (city-level) physical activity policies? [impl2_c1]<br>Yes ☐ [1] [Go to 4.b.] No ☐ [0] I don't know ☐ [99]<br><br>4.b. How would you rate the level of involvement of the national health sector in the implementation stage for developing subnational (city-level) physical activity policies? [impl2a_c1]<br>No involvement ☐[0] ☐[1] ☐[2] ☐[3] ☐[4] ☐[5] Extensive involvement; I don't know ☐[99]<br><br>4.c. (OPTIONAL) – Could you share more information on how the national health sector influences or participates in the implementation stage for the development of subnational (city-level) physical activity policies? [Open Ended Question] [impl3_c1]<br>________________________________________<br>________________________________________<br>4.d. Is there coordination between the national level and the subnational (city-level) level health sectors during the implementation process of subnational (city-level) physical activity policies? [impl4_c1]<br>Yes ☐ [1] No ☐ [0] I don't know ☐ [99] |
| 96 | INTEGRATE | B. Policy operational structure (Subnational city-level policies) | 5. Evaluation stage [eval_c1]<br>Process in which a policy is evaluated to assess its effectiveness concerning the policy objectives. |

| No. | Tool | Indicator / Category | Description |
|---|---|---|---|
| | | | Thinking about the evaluation stage for the development of subnational (city-level) physical activity policies....<br><br>5.a. To the best of your knowledge, are representatives of the national health sector usually participating in the evaluation stage of subnational (city-level) physical activity policies? [eval2_c1]<br>Yes ☐ [1] [Go to 5.b.] No ☐ [0] I don't know ☐ [99]<br><br>5.b. How would you rate the level of involvement of the national health sector in the evaluation stage of subnational (city-level) physical activity policies? [eval2a_c1]<br>No involvement ☐[0] ☐[1] ☐[2] ☐[3] ☐[4] ☐[5] Extensive involvement; I don't know ☐[99]<br><br>5.c. (OPTIONAL) – Could you share more information on how the national health sector influences or participates in the evaluation stage for the development of subnational (city-level) physical activity policies? [Open Ended Question] [eval3_c1]<br>________________________________________________________________<br>________________________________________________________________<br><br>5.d. Is there a formal (e.g., legally binding or a policy statement) process to evaluate the work (priorities, responsibilities, implementation) related to subnational (city-level) physical activity policies between the national level and the subnational (city-level) level health sectors? [eval4_c1]<br>Yes ☐ [1] No ☐ [0] I don't know ☐ [99] |
| 97 | INTEGRATE | B. Policy operational structure (Subnational city-level policies) | 6. To what extent do you consider your office (national health sector) to be involved in agenda setting, policy formulation, adoption (decision making), implementation and evaluation of regional physical activity policies for the following cities? [part_desc_c1] Please review the survey invitation email, which specifies for which cities you are responding to this question.<br>Capital city: No involvement ☐[0] ☐[1] ☐[2] ☐[3] ☐[4] ☐[5] Extensive involvement; I don't know ☐[99]<br>Second city of study: No involvement ☐[0] ☐[1] ☐[2] ☐[3] ☐[4] ☐[5] Extensive involvement; I don't know ☐[99] |

| No. | Tool | Indicator / Category | Description |
|---|---|---|---|
| 98 | INTEGRATE | C. Policy Content | We are now interested in learning about the content of the current national physical activity policy. The following questions relate to the settings, environments, and audiences that are considered relevant for implementing physical activity actions at the national level.<br>1. Does the current physical activity policy document mention the following settings or sectors for implementation? [sett_desc2_c1]<br>Urban environment [urb_c1] ☐ Yes [1] ☐ No [0]<br>Rural environment [rur_c1] ☐ Yes [1] ☐ No [0]<br>Work environment [work_c1] ☐ Yes [1] ☐ No [0]<br>Prison environment [jail_c1] ☐ Yes [1] ☐ No [0]<br>Nurseries and infant schools [nusing_c1] ☐ Yes [1] ☐ No [0]<br>Elementary school [school_c1] ☐ Yes [1] ☐ No [0]<br>High school [highsch_c1] ☐ Yes [1] ☐ No [0]<br>University [college_c1] ☐ Yes [1] ☐ No [0]<br>Health centers, clinics and hospitals [healthcenters_c1] ☐ Yes [1] ☐ No [0]<br>Residential care homes or centers for the elderly [healthsenior_c1] ☐ Yes [1] ☐ No [0]<br>Social care centers [healthsocial_c1] ☐ Yes [1] ☐ No [0]<br>At home [home_c1] ☐ Yes [1] ☐ No [0]<br>Sports and Leisure [sport_c1] ☐ Yes [1] ☐ No [0]<br>Transportation [transp_c1] ☐ Yes [1] ☐ No [0]<br>Tourism [tour_c1] ☐ Yes [1] ☐ No [0]<br>Environment [env_c1] ☐ Yes [1] ☐ No [0]<br>Urban planning [urbp_c1] ☐ Yes [1] ☐ No [0]<br>City [citysett_c1] ☐ Yes [1] ☐ No [0]<br>Neighborhoods [neigh_c1] ☐ Yes [1] ☐ No [0]<br>Other [other_c1] ☐ Yes [1] ☐ No [0] Specify which other setting for implementation: ______ [other_sp_c1] |
| 99 | INTEGRATE | C. Policy Content | We are now interested in knowing if the current national physical activity policy is targeted at specific subgroups of the population. For the following questions, if one or more groups are mentioned as the primary group of interest, as a group of interest for a specific objective, or if implementation or plan is designed for that group, please select it in the table. [subg_desc_c1] |

| No. | Tool | Indicator / Category | Description |
|---|---|---|---|
| | | | 2. List all the target audiences that are mentioned in the current physical activity policy: [subg_desc2_c1]<br>Pre-school children (0-5 years) [subg_presch_c1] ☐ Yes [1] ☐ No [0]<br>Children and/or Adolescents [subg _child_c1] ☐ Yes [1] ☐ No [0]<br>Adults [subg_adult_c1] ☐ Yes [1] ☐ No [0]<br>People with Disabilities [subg_disb_c1] ☐ Yes [1] ☐ No [0]<br>Seniors [subg_senior_c1] ☐ Yes [1] ☐ No [0]<br>People with chronic diseases [subg_cronic_c1] ☐ Yes [1] ☐ No [0]<br>Pregnant women [subg_pregnant_c1] ☐ Yes [1] ☐ No [0]<br>Others [subg_other_c1] ☐ Yes [1] ☐ No [0]<br>Specify the other target audience: [subg _other_sp_c1] ____________ |
| 100 | INTEGRATE | C. Policy Content | 3. Are there other sectors (besides the health sector) mentioned in the national physical activity policy document and that have tangible or concrete responsibilities for implementing the policy? [resp1_c1]<br>☐ No [0]<br>☐ Yes [1]<br><br>3a. Which additional sectors at the national level are mentioned in the current physical activity policy, and have specific implementation responsibilities?<br>[resp2_c1]<br>Sport [resp_sport_c1] ☐ Yes [1] ☐ No [0] ☐ Don't know [99] ☐ N/A [88]<br>Recreation and leisure [resp_rec_c1] ☐ Yes [1] ☐ No [0] ☐ Don't know [99] ☐ N/A [88]<br>Education [resp_edu_c1] ☐ Yes [1] ☐ No [0] ☐ Don't know [99] ☐ N/A [88]<br>Transport [resp_trans_c1] ☐ Yes [1] ☐ No [0] ☐ Don't know [99] ☐ N/A [88]<br>Environment [resp_env_c1] ☐ Yes [1] ☐ No [0] ☐ Don't know [99] ☐ N/A [88]<br>Urban/rural planning and design [resp_urbp_c1] ☐ Yes [1] ☐ No [0] ☐ Don't know [99] ☐ N/A [88]<br>Culture [resp_cult_c1] ☐ Yes [1] ☐ No [0] ☐ Don't know [99] ☐ N/A [88]<br>Tourism [resp_tour_c1] ☐ Yes [1] ☐ No [0] ☐ Don't know [99] ☐ N/A [88] |

| No. | Tool | Indicator / Category | Description |
|---|---|---|---|
| | | | Public finance [resp_pubfin_c1] ☐ Yes [1] ☐ No [0] ☐ Don't know [99] ☐ N/A [88]<br>Work and employment [resp_work_c1] ☐ Yes [1] ☐ No [0] ☐ Don't know [99] ☐ N/A [88]<br>Research [resp_inv_c1] ☐ Yes [1] ☐ No [0] ☐ Don't know [99] ☐ N/A [88]<br>Other sectors [resp_other_c1] ☐ Yes [1] ☐ No [0] ☐ Don't know [99] ☐ N/A [88]<br>Specify other sectors: _____________ [resp_other_sp_c1] |
| 101 | INTEGRATE | D. Policy implementation | The following section aims to understand the implementation stage of the current national level physical activity policies. Policy implementation includes translating statements, ideas, goals, and/or objectives mentioned in the policy documents into practice. For example, a policy document may mention building new facilities as strategies to increase participation in physical activity. Implementation of this statement means having the new facilities built.<br><br>1. Please estimate to what extent the current physical activity policies have been implemented. [implement1_c1]<br>If the policy has been fully implemented, please grade its implementation as 10. Please grade the implementation of a policy from 7 to 9 if most of its statements have been implemented. Please grade the implementation of a policy from 4 to 6 if around half of its statements have been implemented. Please grade the implementation of a policy from 1 to 3 if only a minority of its statements have been implemented. Finally, if the policy has not been implemented at all, please grade it as 0.<br>National-level policy: Not implemented at all ☐[0] ☐[1] ☐[2] ☐[3] ☐[4] ☐[5]☐[6] ☐[7] ☐[8] ☐[9] ☐[10] Fully implemented; Don't know ☐[99]<br><br>1.a. (OPTIONAL) – Could you share more information on why you selected that implementation grade? [Open Ended Question] [implementation_op_c1] |
| 102 | INTEGRATE | D. Policy implementation | 2. Is there funding allocated to the implementation of national physical activity policies in your country?<br>[funding_c1]<br>Yes ☐ [1] [Go to 2.a.] No ☐ [0] Don't know ☐ [99]<br><br>2.a. In addition to the national level, does this funding support the implementation of physical activity policies at other levels? [funding2_c1] |

| No. | Tool | Indicator / Category | Description |
|---|---|---|---|
| | | | a) ☐ Yes, state/regional/province/department level [1]<br>b) ☐ Yes, city level [2]<br>c) ☐ No, this funding only supports implementation at the national level [0] |
| 103 | INTEGRATE | D. Policy implementation | We are now interested in knowing about funding for the implementation of physical activity policies at the national level. [national_fund _c1]<br><br>Type of funding [fund_nat_type_c1]<br>☐ National funds [1]<br>☐ State or regional [2]<br>☐ Subnational (city-level) funds [3]<br>☐ Other [4]:<br>Specify other type of funding for the NATIONAL level: ________ [fund_nat_type_sp_c1]<br><br>Recurrence [fund_nat_recc_c1]<br>☐ Not recurrent (single fund allocation) [1]<br>☐ Each government term (i.e., each<br>government cycle according to the rules of each city/country) [2]<br>- How long does each government term last (i.e., what is the official duration of a regime before there is a change or renewal of government by elections, etc., e.g., 4 years) _____ [fund_nat_period_c1]<br>☐ Other [3]<br>Specify other type of recurrence for funding at the NATIONAL level: ________ [fund_nat_recc_sp_c1]<br><br>Funding source [fund_nat_sour _c1]<br>What is the source of funding at the NATIONAL level? |
| 104 | INTEGRATE | D. Policy implementation | We are now interested in knowing about funding for the implementation of physical activity policies at the regional (state or department) level. [reg_fund _c1] [ only if country_c1 == “Czech Republic”]<br><br>Type of funding [fund_st_type_c1] |

| No. | Tool | Indicator / Category | Description |
|---|---|---|---|
| | | | ☐ National funds [1]<br>☐ State or regional [2]<br>☐ Subnational (city-level) funds [3]<br>☐ Other [4]:<br>Specify other type of funding for the STATE (regional, department, provincial) level: _________ [fund_st_type_sp_c1]<br><br>Recurrence [fund_st_recc_c1]<br>☐ Not recurrent (single fund allocation) [1]<br>☐ Each government term (i.e., each government cycle according to the rules of each city/country) [2]<br>- How long does each government term last (i.e., what is the official duration of a regime before there is a change or renewal of government by elections, etc., e.g., 4 years) ______ [fund_st_period_c1]<br>☐ Other [3]<br>Specify other type of recurrence for funding at the STATE or REGIONAL level: _________ [fund_st_recc_sp_c1]<br><br>Funding source [fund_st_sour _c1]<br>What is the source of funding at the STATE or REGIONAL level? |
| 105 | INTEGRATE | D. Policy implementation | We are now interested in knowing about funding for the implementation of physical activity policies at the subnational level (city level). [local_fund _c1]<br><br>Type of funding [fund_loc_type_c1]<br>☐ National funds [1]<br>☐ State or regional [2]<br>☐ Subnational (city-level) funds [3]<br>☐ Other [4]:<br>Specify other type of funding for the SUBNATIONAL (CITY) level: _________ [fund_loc_type_sp_c1] |

| No. | Tool | Indicator / Category | Description |
|---|---|---|---|
| | | | Recurrence [fund_loc_recc_c1]<br>☐ Not recurrent (single fund allocation) [1]<br>☐ Each government term (i.e., each government cycle according to the rules of each city/country) [2]<br>- How long does each government term last (i.e., what is the official duration of a regime before there is a change or renewal of government by elections, etc., e.g., 4 years) ______ [fund_loc_period_c1]<br>☐ Other [3]<br>Specify other type of recurrence for funding at the SUBNATIONAL (CITY) level: _________ [fund_loc_recc_sp_c1]<br><br>Funding source [fund_loc_sour _c1]<br>What is the source of funding at the SUBNATIONAL (CITY) level? |
| 106 | MOVING | National government supports schools to include physical education in school curricula. | 1. Physical education included in the school curricula<br>a. Legislation and regulations<br>b. Standards<br>c. Programmes<br>d. Guidelines<br><br>2. Hours of physical education in school curricula<br>a. Hours specified<br>b. Hours not specified<br><br>3. Physical education included in the school curricula of primary and secondary schools<br>a. In primary AND secondary schools<br>b. In primary OR secondary school<br><br>4. M&E, enforcement, funding<br>a. Monitoring and evaluation provisions<br>b. Enforcement provisions<br>c. Funding provisions |

| No. | Tool | Indicator / Category | Description |
|---|---|---|---|
| 107 | MOVING | National government supports policies to promote physical activity to children and adolescents during school hours. | 1. Physical activity during school hours<br>a. Mandatory<br>b. Voluntary<br>c. None<br><br>2. Physical activity in the school curricula<br>d. In primary AND secondary schools<br>e. In primary OR secondary school<br><br>3. M&E, enforcement, funding<br>a. Monitoring and evaluation provisions<br>b. Enforcement provisions<br>c. Funding provisions |
| 108 | MOVING | National government supports policies to promote physical activity to children and adolescents outside of school hours. | 1. Physical activity outside school hours<br>a. Mandatory<br>b. Voluntary<br>c. None<br><br>2. Physical activity promotion at school settings<br>a. In primary AND secondary schools<br>b. In primary OR secondary school<br><br>3. M&E, enforcement, funding<br>a. Monitoring and evaluation provisions<br>b. Enforcement provisions<br>c. Funding provisions |
| 109 | MOVING | National government supports community level and mass participation initiatives to promote physical activity. | 1. Community and mass participation initiatives<br>a. Mass participation initiatives and events<br>b. Community level<br><br>2. Focus of initiatives<br>a. Adolescents |

| No. | Tool | Indicator / Category | Description |
|---|---|---|---|
| | | | b. Participants of all abilities<br>c. Least active groups, vulnerable/marginalised people<br><br>3. M&E, enforcement, funding<br>a. Monitoring and evaluation provisions<br>b. Enforcement provisions<br>c. Funding provisions |
| 110 | MOVING | National government supports policies to promote physical activity. | 1. Policies to promote physical activity<br>a. Legislation and regulations<br>b. Standards<br>c. Programmes<br>d. Guidelines<br><br>2. Focus of policies<br>a. Adolescents<br>b. Participants of all abilities<br>c. Least active groups, vulnerable/marginalised people<br><br>3. M&E, enforcement, funding<br>a. Monitoring and evaluation provisions<br>b. Enforcement provisions<br>c. Funding provisions |
| 111 | MOVING | National government supports financial incentives for individuals to promote physical activity. | 1. Focus of financial incentives<br>a. Adolescents<br>b. Participants of all abilities<br>c. Least active groups, vulnerable/marginalised people<br><br>2. M&E, enforcement, funding<br>a. Monitoring and evaluation provisions<br>b. Enforcement provisions<br>c. Funding provisions |

| No. | Tool | Indicator / Category | Description |
|---|---|---|---|
| 112 | MOVING | National government supports the inclusion of physical activity promotion in the pre- and in-service training for non-health care professionals. | 1. Physical activity training for non-health care professionals<br>a. Yes<br>b. No<br><br>2. Physical activity training for non-health care professionals<br>a. Mandatory<br>b. Voluntary<br><br>3. Inclusion of physical activity in the training of non-health care professionals<br>a. For more than one non-health care profession<br>b. For one non-health care profession<br><br>4. Training based on competency-based standards<br>a. Yes<br>b. No<br><br>3. M&E, enforcement, funding<br>a. Monitoring and evaluation provisions<br>b. Enforcement provisions<br>c. Funding provisions |
| 113 | MOVING | National government supports the inclusion of physical activity in the workplace. | 1. Physical activity promotion in the workplace<br>a. Legislation and regulations<br>b. Standards<br>c. Programmes<br>d. Guidelines<br><br>4. M&E, enforcement, funding<br>a. Monitoring and evaluation provisions<br>b. Enforcement provisions<br>c. Funding provisions |

| No. | Tool | Indicator / Category | Description |
|---|---|---|---|
| 114 | MOVING | National government supports design guidelines and/or regulations for buildings that encourage physical activity | 1. Policies for buildings that encourage physical activity<br>a. Legislation and regulations<br>b. Standards<br>c. Guidelines<br><br>2. Active design guidelines for sport facilities<br>a. Yes<br>b. No<br><br>3. Focus of guidelines and/or regulations<br>a. Adolescents<br>b. Participants of all abilities<br>c. Least active groups, vulnerable/marginalised people<br><br>4. M&E, enforcement, funding<br>a. Monitoring and evaluation provisions<br>b. Enforcement provision<br>c. Funding provisions |
| 115 | MOVING | National government supports design guidelines and regulations for the outside of buildings that encourage physical activity. | 1. Policies for outside buildings that encourage physical activity<br>a. Legislation and regulations<br>b. Standards<br>c. Guidelines<br><br>2. Focus of guidelines and/or regulations<br>a. Adolescents<br>b. Participants of all abilities<br>c. Least active groups, vulnerable/marginalised people<br><br>3. M&E, enforcement, funding<br>a. Monitoring and evaluation provisions<br>b. Enforcement provision<br>c. Funding provisions |

| No. | Tool | Indicator / Category | Description |
|---|---|---|---|
| 116 | MOVING | National government supports the facilitation of open/green space that encourages physical activity. | 1. Policies for open/green spaces that encourage physical activity<br>a. Legislation and regulations<br>b. Standards<br>c. Guidelines<br><br>2. Focus of policies<br>a. Adolescents<br>b. Participants of all abilities<br>c. Least active groups, vulnerable/marginalised people<br><br>3. M&E, enforcement, funding<br>a. Monitoring and evaluation provisions<br>b. Enforcement provision<br>c. Funding provisions |
| 117 | MOVING | National government supports the incorporation of walking and cycling infrastructure in urban and rural plans. | 1. Walking and cycling infrastructure policies<br>a. Legislation and regulations<br>b. Standards<br>c. Guidelines<br><br>2. Focus of policies<br>a. Adolescents<br>b. Participants of all abilities<br>c. Least active groups, vulnerable/marginalised people<br><br>3. M&E, enforcement, funding<br>d. Monitoring and evaluation provisions<br>e. Enforcement provision<br>f. Funding provisions |
| 118 | MOVING | National government supports prioritising integrated urban design and mixed land-use policies | 1. Urban design and land-use policies<br>a. Legislation and regulations<br>b. Standards<br>c. Guidelines |

| No. | Tool | Indicator / Category | Description |
|---|---|---|---|
| | | prioritising compact, mixed-land use in urban and rural plans. | 2. Focus of policies<br>a. Adolescents<br>b. Participants of all abilities<br>c. Least active groups, vulnerable/marginalised people<br><br>3. M&E, enforcement, funding<br>a. Monitoring and evaluation provisions<br>b. Enforcement provision<br>c. Funding provisions |
| 119 | MOVING | National government supports increasing access to public open space and green spaces in urban and rural plans. | 1. Open space and green spaces policies<br>a. Legislation and regulations<br>b. Standards<br>c. Guidelines<br><br>2. Focus of policies<br>a. Adolescents<br>b. Participants of all abilities<br>c. Least active groups, vulnerable/marginalised people<br><br>3. M&E, enforcement, funding<br>a. Monitoring and evaluation provisions<br>b. Enforcement provision<br>c. Funding provisions |
| 120 | MOVING | National government supports the increased provision of public transport. | 1. Public transport policies<br>a. Legislation and regulations<br>b. Standards<br>c. Programmes<br>d. Guidelines<br><br>2. Focus of policies<br>a. Adolescents |

| No. | Tool | Indicator / Category | Description |
|---|---|---|---|
| | | | b. Participants of all abilities<br>c. Least active groups, vulnerable/marginalised people<br><br>3. M&E, enforcement, funding<br>a. Monitoring and evaluation provisions<br>b. Enforcement provision<br>c. Funding provisions |
| 121 | MOVING | National government supports increasing road safety actions to protect pedestrians, cyclists etc. | 1. Road safety polices<br>a. Legislation and regulations<br>b. Standards<br>c. Programmes<br>d. Guidelines<br><br>2. Focus of policies<br>a. Adolescents<br>b. Participants of all abilities<br>c. Least active groups, vulnerable/marginalised people<br><br>3. M&E, enforcement, funding<br>a. Monitoring and evaluation provisions<br>b. Enforcement provision<br>c. Funding provisions |
| 122 | MOVING | National government supports a public information campaign that promotes transport. | 1. Aim of the campaign<br>a. increase awareness about road safety<br>b. promote the use of public transport<br>c. promote active transport<br><br>2. Campaign includes social marketing<br>a. Yes<br>b. No<br><br>3. Focus of the campaign |

| No. | Tool | Indicator / Category | Description |
|---|---|---|---|
| | | | a. Inactive population segments<br>b. Adolescents<br>c. Adults<br>d. Entire population<br><br>4. Campaign signposts to services or more information<br>a. Yes<br>b. No<br><br>5. M&E, enforcement, funding<br>a. Monitoring and evaluation provisions<br>b. Enforcement provision<br>c. Funding provisions |
| 123 | MOVING | National government supports active transport. | 1. Policies that promote active transport<br>a. To and from school<br>b. To and from work<br>c. Active transport in general<br><br>2. Active transport policies<br>a. Legislation and regulations<br>b. Standards<br>c. Programmes<br>d. Guidelines<br><br>3. Active transport through programmes<br>a. Multiple programmes<br>b. One programme<br><br>4. Active transport to and from school<br>a. In primary AND secondary schools<br>b. In primary OR secondary school |

| No. | Tool | Indicator / Category | Description |
|---|---|---|---|
| | | | 5. M&E, enforcement, funding<br>a. Monitoring and evaluation provisions<br>b. Enforcement provision<br>c. Funding provisions |
| 124 | MOVING | National government supports public awareness, mass media and informational campaigns and social marketing promoting physical activity. | 1. Mass communication campaign includes social marketing<br>a. Yes<br>b. No<br><br>2. Focus of the campaign<br>a. Inactive population segments<br>b. Adolescents<br>c. Adults<br>d. Entire population<br><br>4. Campaign signposts to services or more information<br>a. Yes<br>b. No<br><br>5. M&E, enforcement, funding<br>1. Monitoring and evaluation provisions<br>2. Enforcement provision<br>3. Funding provisions |
| 125 | MOVING | National government develops and communicates physical activity guidelines. | 1. Target of physical activity guidelines<br>a. young children and adolescents<br>b. general population<br><br>2. Dissemination of physical activity guidelines<br>a. Through mass communication campaigns targeted at youth<br>b. Through mass communication campaigns<br>c. No dissemination through mass communication campaigns |

| No. | Tool | Indicator / Category | Description |
|---|---|---|---|
| | | | 3. Campaigns signpost to services or more information<br>a. Yes, and has concurrent policy actions, programs or environmental changes to support the behaviours targeted<br>b. Yes<br>c. Camping stands alone/ no signposting<br><br>4. M&E, enforcement, funding<br>a. Monitoring and evaluation provisions<br>b. Enforcement provision<br>c. Funding provisions |
| 126 | MOVING | National government supports the inclusion physical activity promotion in the pre- and in-service training for health professionals | 1. Physical activity training for health care professionals<br>a. Mandatory<br>b. Voluntary<br><br>2. Inclusion of physical activity in the training of health professionals<br>a. For one health care profession<br>b. For more than one health profession<br><br>3. Training based on standards<br>c. Yes<br>d. No<br><br>4. M&E, enforcement, funding<br>a. Monitoring and evaluation provisions<br>b. Enforcement provisions<br>c. Funding provisions |
| 127 | MOVING | National government supports the inclusion of physical activity counselling, | 1. Physical activity counselling, assessment and physical activity prescriptions<br>a. Legislation and regulations<br>b. Standards |

| No. | Tool | Indicator / Category | Description |
|---|---|---|---|
| | | assessment and physical activity prescriptions in primary care. | c. Programmes<br>d. Guidelines<br><br>2. Measures include advice and counselling for<br>a. General public<br>b. Children and adolescents<br>c. Children and adolescents with obesity-related issues<br><br>3. M&E, enforcement, funding<br>a. Monitoring and evaluation provisions<br>b. Enforcement provisions<br>c. Funding provisions |
| 128 | MOVING | National government supports the inclusion of physical activity counselling, assessment and physical activity prescriptions in health care and outpatient settings. | 1. Physical activity counselling, assessment and physical activity prescriptions<br>a. Legislation and regulations<br>b. Standards<br>c. Programmes<br>d. Guidelines<br><br>2. Measures include advice and counselling for<br>a. General public<br>b. Children and adolescents<br>c. Children and adolescents with obesity-related issues<br><br>3. M&E, enforcement, funding<br>a. Monitoring and evaluation provisions<br>b. Enforcement provisions<br>c. Funding provisions |
| 129 | PA-EPI | E01 | Evidence informed, quality mandatory physical education that promotes and supports the ideals of equity, diversity and inclusion and adheres to defined standards is part of the curricula in all schools. |

| No. | Tool | Indicator / Category | Description |
|---|---|---|---|
| 130 | PA-EPI | E02 | National and/or subnational initiatives are in place to promote and support school-related physical activity both at school and in other settings. These initiatives should employ an inter-sectoral approach and collaborative multi-agency partnerships (e.g., links with out-of-school sports clubs, active breaks/recess, walking clubs). |
| 131 | PA-EPI | E03 | There are shared use agreements that utilise school spaces. Community access is supported by initiatives to promote and support opportunities for physical activity for all persons outside of normal school hours. |
| 132 | PA-EPI | E04 | National and/or sub-national policies are in place to promote and support safe active travel to and from school. |
| 133 | PA-EPI | T01 | Regulations are in place that provide a variety of infrastructures to support safe walking and/or cycling and/or wheeling, including measures to calm speed, reduce vehicle traffic and enhance active mobility. |
| 134 | PA-EPI | T02 | There is a funded implementation plan, led by the appropriate level/s of government, to achieve improvements in active travel and increased use of public transport. |
| 135 | PA-EPI | T03 | Guidelines and tools to support infrastructure for active mobility and/or transport plans and systems that encourage physical activity are promoted and disseminated. |
| 136 | PA-EPI | UD01 | Policies or regulations that take a “health in all” approach is adopted to reallocate space from motorised transport to active travel and/or recreation purposes. |
| 137 | PA-EPI | UD02 | Governments adopt land use policies, and planning processes, consistent with principles of mixed land use, compact urban design, and/or provision of green open spaces to support physical activity and reduce motorised transport. |
| 138 | PA-EPI | UD03 | There are guidelines and/or regulations that improve universal and equitable access to safe outdoor and indoor spaces and facilities where people can be physically active. |
| 139 | PA-EPI | H01 | Guidelines and regulations in healthcare include routine screening for physical activity and, for all insufficiently active patients, brief advice, and referral to appropriately trained practitioners and/or physical activity opportunities. |
| 140 | PA-EPI | H02 | There are consistent policies for promoting and supporting physical activity in primary and secondary healthcare settings among at-risk groups, such as people with type 2 diabetes and older adults (e.g., protocols for the assessment of the physical activity capacity; accessible, affordable, and tailored physical activity programmes; and training for caregivers for delivering physical activity programmes within residential aged care). |

| No. | Tool | Indicator / Category | Description |
|---|---|---|---|
| 141 | PA-EPI | MM01 | There are national and/or subnational public policies in place that ensure media and education campaigns that promote and support physical activity are sustained and monitored (e.g., by making them part of, or aligning them with, a national action plan on physical activity and the physical activity guidelines). |
| 142 | PA-EPI | MM02 | There are clear, consistent policies to ensure that multiple media modes/channels (e.g., via posters, social media, radio as well as TV) combined with complementary community initiatives are used to promote the benefits of physical activity and disseminate guidelines which align with the WHO physical activity recommendations. |
| 143 | PA-EPI | C01 | Public policies are in place to support the implementation of whole-of-community approaches to promote physical activity and networking to strengthen resources and exchange experiences (e.g., WHO Healthy Cities, Active Cities, Partnerships for Healthy Cities). |
| 144 | PA-EPI | C02 | There are public policies in place to foster partnerships for shared use of public spaces and facilities for community-based and community-led physical activity programmes. |
| 145 | PA-EPI | SP01 | There are national and/or subnational evidence informed 'Sport and Recreation for All' policies that prioritise investment in initiatives that target the least active, as well as disadvantaged groups. |
| 146 | PA-EPI | SP02 | There are national and/or subnational evidence informed policies or action plans in place that ensure equitable access to sport and recreation spaces and places for all. |
| 147 | PA-EPI | SP03 | There is government support for programs designed to encourage sports clubs to promote health-enhancing physical activity and other health behaviours (e.g., 'sports clubs for health' and 'health promoting sport clubs'). |
| 148 | PA-EPI | W01 | There are national and/or sub-national policy initiatives and infrastructure development programmes in place to promote and support safe active travel to and from the workplace. |
| 149 | PA-EPI | W02 | There are concepts and regulations for buildings, plots and the environment in place that promote and support employers to create physically active workplace environments through building design and provision of adequate facilities (both indoor and outdoor). |
| 150 | PA-EPI | L01 | There is strong, visible, political support (at the head of state/cabinet level) for creating health-promoting policy environments to improve population levels of physical activity and reduce inactivity related non-communicable diseases and their related inequalities. Political responsibility for health-related physical activity is clearly allocated within the governmental structures. |

| No. | Tool | Indicator / Category | Description |
|---|---|---|---|
| 151 | PA-EPI | L02 | There is a comprehensive up-to-date plan (including timeline, targets, funding, priority policy and programme strategies) linked to national needs and priorities to increase population physical activity. |
| 152 | PA-EPI | L03 | Priorities are given to reduce inequalities in relation to inactivity related non-communicable diseases in the comprehensive plan (above). |
| 153 | PA-EPI | L04 | There are clearly defined, evidenced informed population physical activity guidelines for all age groups and for people living with non-communicable diseases, pregnant women, and people with disabilities. |
| 154 | PA-EPI | G01 | There are reliable procedures to restrict commercial influences related to physical activity environments where there are conflicts of interest with improving population physical activity levels (e.g., restricting lobbying influences that limit physical activity opportunities). |
| 155 | PA-EPI | G02 | There are procedures in place for using evidence in the development of physical activity policies. |
| 156 | PA-EPI | G03 | The government ensures access to and regular dissemination of physical activity guidelines and key documents to the public. |
| 157 | PA-EPI | G04 | The government fosters the cooperation and coordination of all sectors to align with strategic plans to improve the physical activity environment, and where appropriate, promotes civil society participation to develop and implement these plans. |
| 158 | PA-EPI | MI01 | There is regular monitoring of physical activity levels across the life-course based on representative samples, against guidelines/standards/targets. |
| 159 | PA-EPI | MI02 | There is regular monitoring of physical activity environments across all 8 policy domains (e.g., walkability, built environment). |
| 160 | PA-EPI | MI03 | Physical activity monitoring is systematically linked to the regular monitoring of non-communicable diseases and their related inequalities. |
| 161 | PA-EPI | MI04 | There is regular research and evaluation of policies and major programmes to assess their effectiveness, process, and impact on achieving the goals of the physical activity and health plans. |
| 162 | PA-EPI | MI05 | Progress towards reducing health inequalities related to social and economic determinants of physical activity is regularly monitored. |
| 163 | PA-EPI | FR01 | The budget spent on physical activity promotion across all policy domains is clearly identified and periodically monitored. |

| No. | Tool | Indicator / Category | Description |
|---|---|---|---|
| 164 | PA-EPI | FR02 | There is a sufficient proportion of total health spending assigned to population physical activity promotion. |
| 165 | PA-EPI | FR03 | A sufficient proportion of total research spending is assigned to population physical activity promotion. |
| 166 | PA-EPI | FR04 | A secure funding stream is available for at least one statutory health promotion agency with an objective to improve population physical activity. |
| 167 | PA-EPI | PI01 | There are robust coordination mechanisms across departments and levels of government to ensure policy coherence, alignment and integration of physical activity, and inactivity related non-communicable disease prevention policies across governments. |
| 168 | PA-EPI | PI02 | There are structures and mechanisms for regular, meaningful, and inclusive interactions between government and civil society (academia, professional organizations, public-interest, non-governmental organisations, and citizens) on physical activity policies and other strategies to improve population physical activity and health. |
| 169 | PA-EPI | WD01 | To address the challenge of population physical inactivity, there are sufficient resources and people with necessary skills within the government's workforce (across all 8 policy domains). |
| 170 | PA-EPI | WD02 | Opportunities for training and professional development are provided to relevant individuals across multiple sectors (e.g., the 8 'Policy' domains) regarding the fundamentals of physical activity, its role in public health, and effective strategies for physical activity promotion. |
| 171 | PA-EPI | WD03 | Support and training systems are in place for relevant professionals (e.g., guidelines, toolkits, training workshops/modules/courses). To ensure uptake, accrediting agencies for professional education, and professional licensing entities should include minimum requirements for initial and continuing education in this domain. |
| 172 | PA-EPI | HIAP01 | There are processes in place to ensure that population physical activity and related health outcomes are explicitly and transparently considered and prioritised in the development of all government policies. |
| 173 | PA-EPI | HIAP02 | There are processes (e.g., health impact assessments) to assess and consider health impacts during the development of policies indirectly related to physical activity. |
| 174 | WHO | National communication campaigns on physical activity (1.1.1) | % of countries that have implemented national community-wide public education and awareness campaigns on physical activity in the past 2 years |
| 175 | WHO | National communication campaigns on physical activity with integrated | % of countries that have implemented national physical activity campaigns for physical activity with community links |

| No. | Tool | Indicator / Category | Description |
|---|---|---|---|
| | | links to community-based initiatives (1.1.2) | |
| 176 | WHO | National communication campaigns on physical activity supported by environmental changes (1.1.3) | % of countries that have implemented national physical activity campaigns for physical activity which includes supportive environment links |
| 177 | WHO | National physical activity communication campaigns promoting on co-benefits of physical activity (1.2.1) | % of countries which have conducted a public education and awareness campaign focused on promoting the co-benefits of physical activity |
| 178 | WHO | National mass participation events on physical activity (1.3.1) | % of countries which have conducted at least one free mass participation event on physical activity |
| 179 | WHO | National policy on walking and cycling (2.2.1) | % of countries with national policy on walking and/or cycling |
| 180 | WHO | National policy on public transport (2.2.2) | % of countries with national policies and investment in increasing access to public transport |
| 181 | WHO | National road design standards (2.2.3) | % of countries with design standards for: managing speed where pedestrians and cyclists are present; safe crossings for pedestrians and cyclists; and separation of pedestrians and cyclists from vehicular traffic |
| 182 | WHO | National road safety strategy (2.2.4) | % of countries with national funded road safety strategy |
| 183 | WHO | Road safety assessment on existing road networks (2.2.5) | % of countries with road safety star rating [or safety rating] assessments for existing road networks |
| 184 | WHO | Road safety assessment of new road infrastructure projects (2.2.6) | % of countries with road safety audit [star/safety rating] assessment prior to construction in the design or plans of new road infrastructure |
| 185 | WHO | Legislation on speed limits meeting best practice (2.3.1) | % of countries with the national/ provincial/state speed legislations met the best practice criteria |
| 186 | WHO | Legislation on drink-driving meeting best practice (2.3.3) | % of countries with national/provincial/state drink driving legislations met the best practice criteria |
| 187 | WHO | Legislation on distracted driving (mobile phone use) (2.3.5) | % of countries with the national/provincial/state legislation on distracted driving |
| 188 | WHO | Legislation on distracted driving (drug use) (2.3.5) | % of countries with the national/provincial/state legislation on distracted driving |

| No. | Tool | Indicator / Category | Description |
|---|---|---|---|
| 189 | WHO | National protocols/standards for the management of physical inactivity through primary care (3.2.1) | % countries with national guidelines/protocols/standards for management of physical inactivity in primary health care |
| 190 | WHO | Implementing national policies to promote physical activity in childcare settings (3.3.1) | % of countries implementing national policies promoting population physical activity in childcare settings |
| 191 | WHO | Implementing national policies to promote physical activity in the workplace (3.3.2) | % of countries implementing national policies on physical activity in the workplace |
| 192 | WHO | Implementing national policies to promote community-based physical activity and sports initiatives (3.3.3) | % of countries implementing national policies on community-based physical activity and sports initiatives |
| 193 | WHO | Implementing national policies to promote physical activity in public open spaces (including parks) (3.3.4) | % of countries implementing national policies to promote physical activity in public open spaces (including parks) |
| 194 | WHO | Implementing national policies to promote walking and cycling (3.3.5) | % of countries implementing national policies to increase walking and cycling |
| 195 | WHO | Implementing national policies to promote physical activity as part of active aging (3.4.1) | % of countries implementing national policies on physical activity as part of active aging |
| 196 | WHO | National mHealth initiative (4.3.1) | % of countries applying mHealth in NCD prevention and management |
| 197 | WHO | Operational national NCD policy which includes physical activity (4.1.1) | % of countries with operational national NCD policy, strategy and/or action plan that includes physical activity |
| 198 | WHO | Operational national physical activity policy, strategy, or action plan (4.1.2) | % of countries with operational national PA policy, strategy or action plan on physical activity |
| 199 | WHO | National guidelines on physical activity (4.1.3) | % of countries with national guidelines on physical inactivity in children aged <5, adolescents, adults, and older adults |
| 200 | WHO | National physical activity target (4.1.4) | % of countries with national target(s) for physical activity |

| No. | Tool | Indicator / Category | Description |
|---|---|---|---|
| 201 | WHO | National coordination mechanism for NCDs (4.1.5) | % of countries with present and operational NCD multisectoral commission, agency or mechanism |
| 202 | WHO | National surveillance of physical activity (4.2.1) | % of countries with national surveillance on physical inactivity in children, adolescents and adults |
